# Metagenomics Combined with Activity-Based Proteomics Point to Gut Bacterial Enzymes that Reactivate Mycophenolate

**DOI:** 10.1101/2022.05.19.22274720

**Authors:** Joshua B. Simpson, Josh J. Sekela, Amanda L. Graboski, Valentina B. Borlandelli, Natalie K. Barker, Alicia A. Sorgen, Marissa M. Bivins, Angie L. Mordant, Rebecca L. Johnson, Aadra P. Bhatt, Anthony A. Fodor, Laura E. Herring, Hermen Overkleeft, John R. Lee, Matthew. R. Redinbo

## Abstract

Mycophenolate Mofetil (MMF) is an important immunosuppressant prodrug prescribed to prevent organ transplant rejection and to treat autoimmune diseases. MMF usage, however, is limited by severe gastrointestinal toxicity that is observed in approximately 45% of MMF recipients. The active form of the drug, mycophenolic acid (MPA), undergoes extensive enterohepatic recirculation by bacterial β-glucuronidase (GUS) enzymes, which reactivate MPA from mycophenolate glucuronide (MPAG) within the gastrointestinal tract. GUS enzymes demonstrate distinct substrate preferences based on their structural features, and gut microbial GUS enzymes that reactivate MPA have not been identified. Here, we compare the fecal microbiomes of transplant recipients receiving MMF to healthy individuals using shotgun metagenomic sequencing. We find that neither microbial composition nor the presence of specific structural classes of GUS genes are sufficient to explain the differences in MPA reactivation measured between fecal samples from the two cohorts. We next employed a GUS-specific activity-based chemical probe and targeted metaproteomics to identify and quantify the GUS proteins present in the human fecal samples. The identification of specific GUS enzymes was improved by using the metagenomics data collected from the fecal samples. We found that the presence of GUS enzymes that bind the flavin mononucleotide (FMN) is significantly correlated with efficient MPA reactivation. Furthermore, structural analysis identified motifs unique to these FMN-binding GUS enzymes that provide molecular support for their ability to process this drug glucuronide. These results indicate that FMN-binding GUS enzymes may be responsible for reactivation of MPA and could be a driving force behind MPA-induced GI toxicity.

## Introduction

The prodrug Mycophenolate Mofetil (MMF; CellCept) was approved by the FDA in 1995 and is now widely prescribed to prevent organ transplant rejection and to treat autoimmune diseases.^1, 2^ Mycophenolic Acid (MPA), the active form of MMF, inhibits the lymphocyte isoform of inosine monophosphate dehydrogenase (IMPDH), thereby arresting the proliferation of B and T lymphocytes by limiting their synthesis of guanosine triphosphate (GTP).^2, 3^ Treatment with MMF, however, is often limited by severe GI side effects that are observed in approximately 45% of recipients.^4, 5^

Recent studies have demonstrated that treatment with MMF initiates a shift in the microbiome, and that these changes may mediate drug-associated GI toxicity.^6–8^ Indeed, GI toxicity was dramatically reduced in germ-free mice compared to controls when treated with MMF.^6^ Additionally, treatment with the antibiotic vancomycin was shown to ameliorate GI toxicity in mice administered MMF, establishing the microbiota’s involvement in this unwanted side effect.^8^ When a human subject was administered antibiotics alongside MMF, however, GI toxicity was not alleviated.^7^ Given that organ transplant recipients are typically administered MMF over extended periods, an antibiotic-directed approach is likely not a practical therapeutic approach for severe GI toxicity, as the microbiota also play critical roles in metabolism and homeostasis.^5, 9–13^

MMF is ester-linked with a morpholino ethyl moiety that increases efficacy and bioavailability; this group is removed by host esterases to produce active MPA, which interacts with the microbiota through so-called phase IV metabolism (Supplemental Figure 1).^1, 14, 15^ MPA is glucuronidated in the liver by uridine diphosphate (UDP)-glucuronosyltransferase enzymes to form inactive mycophenolate glucuronide (MPAG), which is excreted via the biliary ducts to the gastrointestinal tract.^15^ Once in the intestines, MPA can be reactivated by bacterial β-glucuronidase (GUS) enzymes (Figure 1). Indeed, mice that received both MMF and vancomycin showed decreased fecal levels of active MPA and higher fecal levels of inactive MPAG compared to mice only receiving MMF, suggesting that gut microbes extensively metabolize MPAG.^8^ MPA that is reactivated in the gut can be reabsorbed into systemic recirculation, altering therapeutic levels of this critical immunosuppressant and exacerbate neutropenia, which is an indicator of excessive neurosuppression.^8, 16^ Within the gut, MPA reduces the integrity of the GI epithelial barrier, resulting in side effects such as vomiting, diarrhea, ulcers, and decreased concentrations of short-chain fatty acids.^5, 17, 18^ Thus, gut microbial GUS enzymes play a crucial role in the many deleterious side effects of MMF.

**Figure 1.**
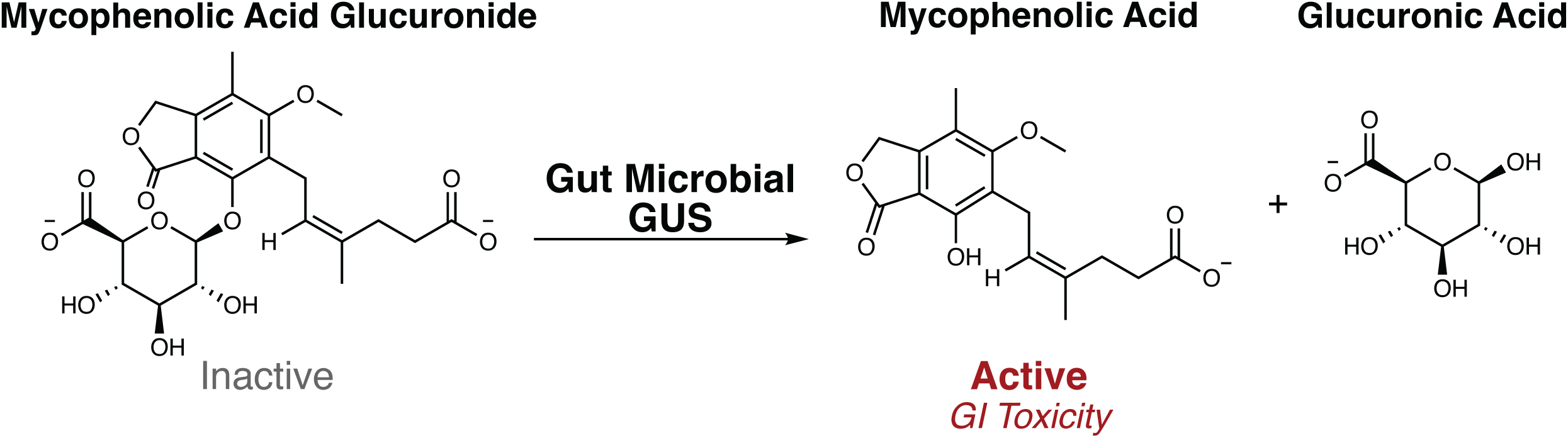
Gut microbial β-glucuronidase (GUS) enzymes remove the glucuronide as a source of carbon, and active MPA is reabsorbed in the gut lumen, contributing to gastrointestinal toxicity.

GUS enzymes reactivate a variety of xenobiotic and endobiotic glucuronides from their inactive glucuronide metabolites, including cancer drugs irinotecan and regorafenib, several NSAIDs, the consumer antimicrobial compound triclosan, as well as several forms of estrogen.^19–23^ Prior metagenomic analyses have identified hundreds of putative GUS enzymes that can be categorized into one of eight unique structural classes displaying distinct substrate preferences.^24–26^ The development of a GUS-specific activity-based chemical probe has enabled the use of targeted metaproteomics to quantify the GUS proteins present and active in human fecal samples.^23, 27^ This technology has been further adapted to identify the structural classes of GUS enzymes that are responsible for reactivation of irinotecan and triclosan.^21, 23^ While gut microbial GUS enzymes likely reactivate MPA, the structural details of this reactivation reaction have not been explored towards identifying which of the hundreds of potential GUS orthologs may be primarily responsible. We hypothesized that bacterial GUS abundance and composition would vary between transplant recipients receiving MMF and healthy individuals, and that these differences would enable us to pinpoint the specific GUS enzymes responsible for MPA reactivation. Here, we compare the microbiomes and metagenomic GUS profiles of transplant recipients receiving MMF to healthy individuals using shotgun metagenomic sequencing. We then analyze the fecal samples using GUS-targeted metaproteomics leveraging the cohort’s metagenomics data as a reference. The results obtained show that, while metagenomic sequencing data are insufficient to show correlations with MPA reactivation rates, the metaproteomics data revealed clear associations with the levels of FMN-binding GUS proteins and the rates of drug reactivation in individual samples. Finally, we explore the potential structural basis of these observations. Together, these data suggest for the first time that specific FMN-binding GUS enzymes may be responsible for MPA-induced GI toxicity.

## Results

### MMF Treatment and Gut Microbial Composition

We collected fecal samples from five renal transplant recipients receiving MMF and four healthy individuals. The transplant recipients had ages ranging from their 40s to their 70s, and all were receiving a calcineurin inhibitor-based maintenance regimen as well as MMF. Further details of the transplant recipients are found in Supplemental Table 1. In four cases, the transplant recipient fecal samples were collected closely following transplantation (days 4-9), but in one case (T3) it was collected 227 days after the transplant. The healthy individuals had ages ranging from their 20s to their 50s and had not received antibiotics for several months prior to fecal collection. All individuals, both transplant recipients and controls, were male.

We first examined the fecal metagenomic profiles using shotgun metagenomic sequencing (Supplemental Figure 2). Results at the class level of taxonomies are shown in Figure 2A, while phyla, order, family, genus, and species levels of taxonomies are shown in Supplemental Figures 3-7, respectively. While all samples contained microbes of the *Clostridia* class, distinct class differences were observed between groups. Specifically, the flora of transplant recipients who received MMF had the presence of *Bacilli*, *Gammaproteobacteria*, and *Erysipelotrichia*, which were not observed in healthy individual samples; in contrast, healthy individuals uniquely had the presence of *Actinobacteria* and *Verrucomicrobiae* (Figure 2A).

**Figure 2.**
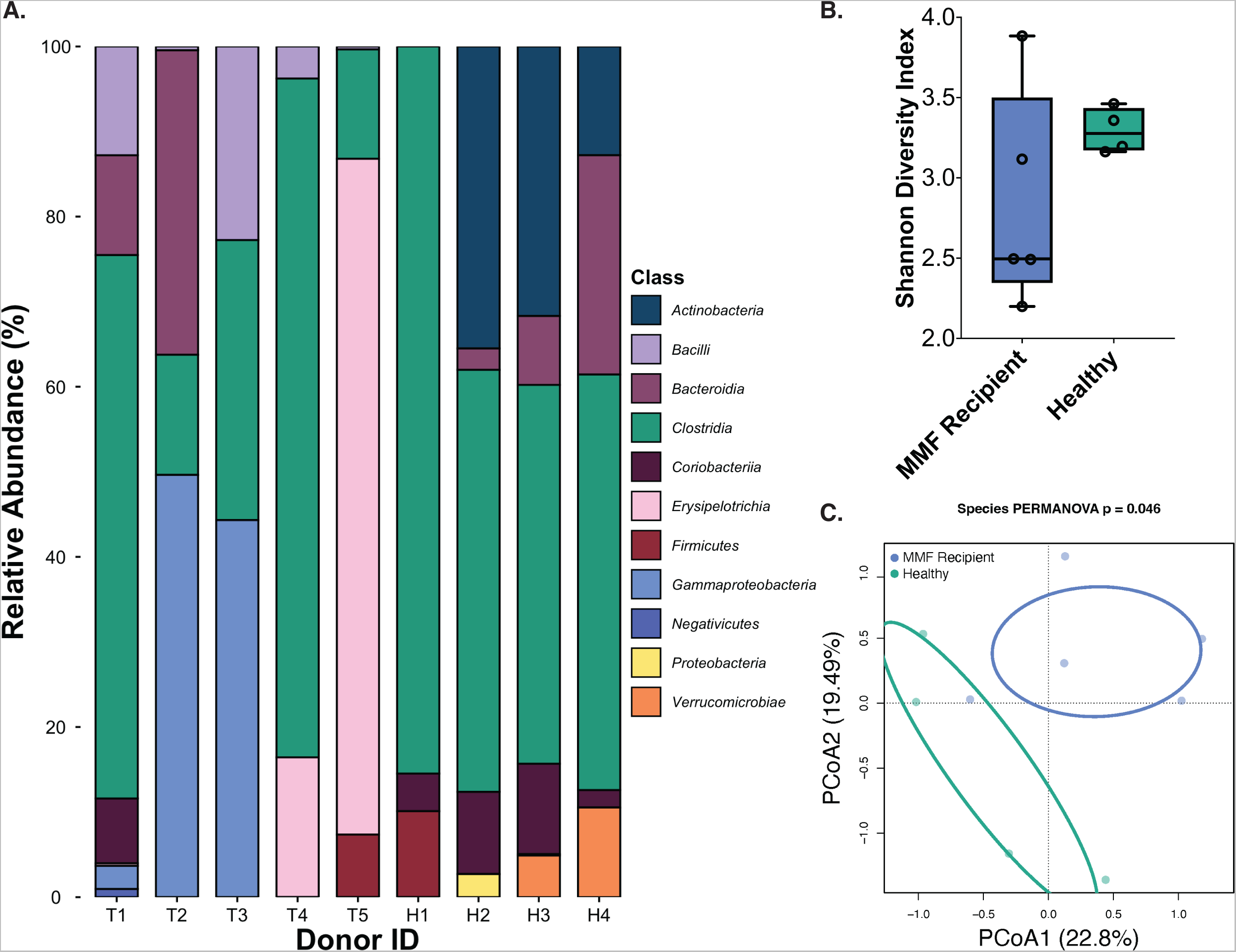
Metagenomic Shotgun Sequencing profiles for MMF recipients (blue; T1-T5) and healthy individuals (green; H1-H4). (A) Relative abundance of intestinal bacteria by Class. (B) Alpha diversity by the Shannon Diversity Index. (C) Bray Curtis PCoA ordination of Beta diversity at the species level from shotgun metagenomic sequencing and shown with PC1 and PC2 components.

**Figure 3.**
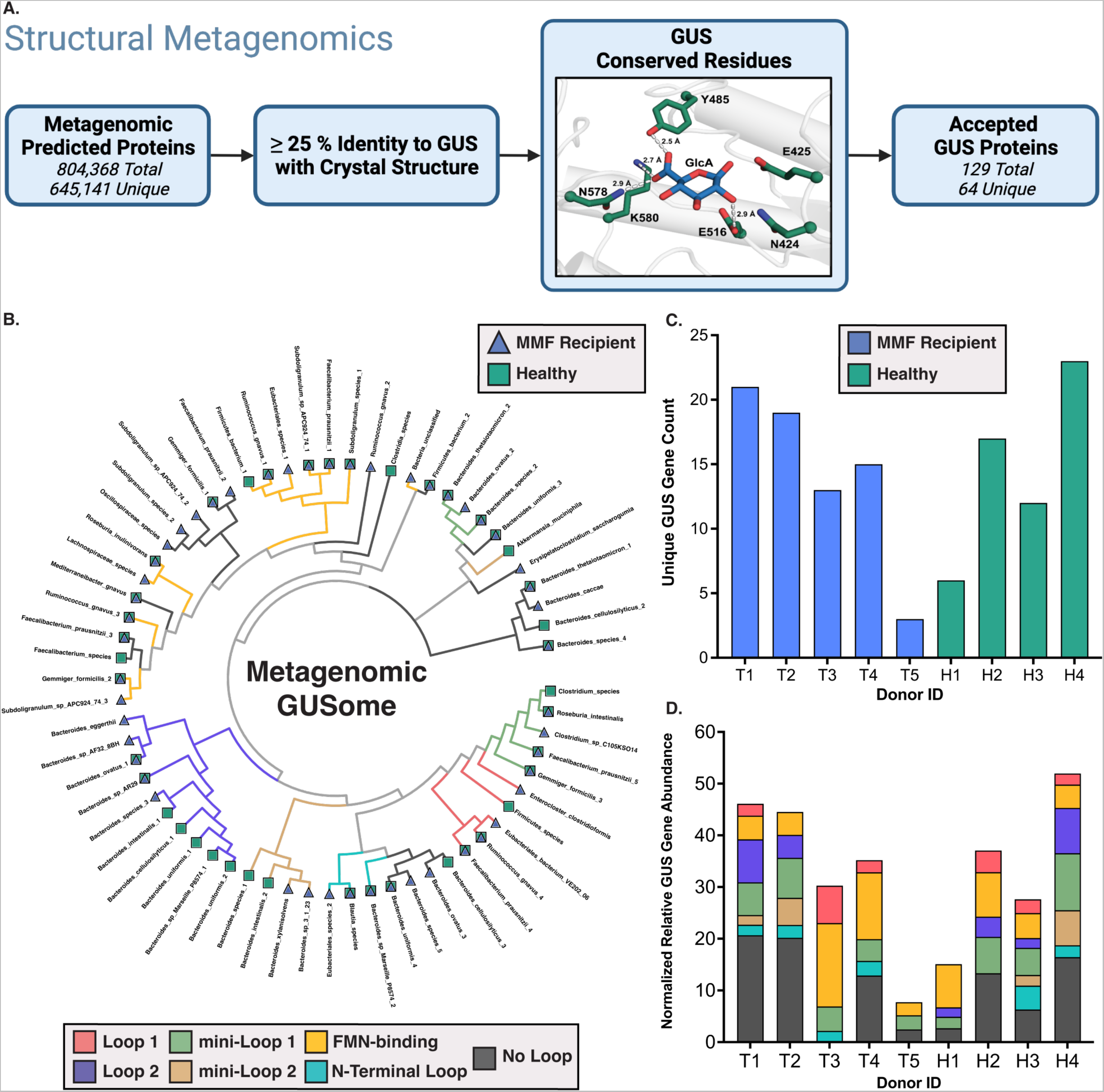
(A) Structural metagenomics workflow for identification of GUS proteins. (B-D) Metagenomic GUS gene profiles for MMF recipients (blue; T1-T5) and healthy individuals (green; H1-H4). (B) Cladogram reflecting non-redundant GUS gene sequences across the cohort. Each node represents GUS protein sequences with ≥ 90 % identity. GUS class and treatment groups from which genes were derived are indicated. Unique GUS gene counts for each donor (C) and Normalized Gene Abundances clustered by GUS class (D) are also shown. Figure 3A was created with BioRender.com.

There was similar alpha diversity between the fecal samples from MMF-treated transplant recipients and those from healthy individuals as measured by the Shannon Diversity Index (Figure 2B). However, the beta diversity at the species level between transplant recipients and healthy individuals was different (Figure 2C; p = 0.046).

### Stool Metagenomic GUSome Profiles Do Not Differ Between Kidney Transplant Recipients Who Received MMF and Healthy Individuals

The gene catalogs extrapolated from shotgun metagenomics for our cohort contained 884,299 total translated protein sequences, and we employed a structure-guided approach to identify those that encoded genes for GUS enzymes (Figure 3A; Supplemental Figure 2). Each sequence was aligned to the 17 representative gut microbial GUS enzymes with reported crystal structures; sequences with ≥25% identity to at least one representative GUS enzyme were then assessed for the presence of 7 conserved active site residues essential for glucuronide hydrolysis.^24^ Predicted protein sequences that met both the identity threshold and contained all conserved residues were accepted as GUS enzymes. Taxonomy of origin and GUS structural class for each sequence were then assigned by comparing protein sequences to the RefSeq Select database. In total, 130 genes for GUS proteins were detected, and clustering into groups with ≥90% sequence identity collapsed these into a final collection of 64 GUS proteins (a “GUSome” for this dataset; Figure 3B). Nearly all the sequences are derived from either *Bacteroidetes* or *Firmicutes* phyla (Supplemental Figure 8).

We then turned to the distinct structural clades that have been assigned to gut microbial GUS enzymes.^24–26^ Several clusters within the metagenomic GUSome belonging to a structural class were found to be either uniquely present or notably missing from the fecal samples taken from transplant recipients receiving MMF. Notably, *Oscillospiraceae*-derived “No Loop” GUS genes are only found within MMF recipients, while genes for *Bacteroidaceae*-derived “Loop 2” GUS enzymes are entirely absent from this group (Figure 3B). There were other instances of individual GUS genes within a structural class being present within only one treatment group. However, we observed no differences between MMF recipients and healthy individuals in overall GUS gene abundance overall or for any structural class (Supplemental Figure 9).

The number of unique GUS genes ranged from 3 to 21 in the samples from MMF-treated transplant recipients, and 6 to 23 in samples from healthy individuals (Figure 3C). These ranges are akin to those observed earlier in the 139 donor samples from the Human Microbiome Project stool sample database, which showed a range of 4-41 unique GUS proteins per individual (and termed an individual’s “GUSome”).^24^ Similarly, the individual GUSomes of each sample in the current study exhibited a range of gene abundances for GUS enzymes of different structural classes (Figure 3D). While the samples from MMF-treated transplant recipient T5 contained genes from only three GUS classes, several samples contained genes for all seven classes (T1, H3, H4) or six of seven (T2). Together, these results suggest that the metagenomic profiles of the MMF-treated transplant recipients and healthy individuals contain genes coding a similar range of structurally diverse GUS proteins.

### Differences in Mycophenolate Reactivation Between Fecal Samples

Given that shotgun metagenomics reflect only the genes present in the microbiome and not necessarily the proteins that are expressed and active, we sought to explore the potential differences in bulk GUS enzyme activity toward mycophenolate-glucuronide hydrolysis between fecal samples collected from the five renal transplant recipients receiving MMF and the four healthy individuals. To do so, we extracted the complex protein lysates from each fecal sample and measured the rate of reactivation of MPA from the inactive metabolite MPAG. This approach has been employed previously to examine drug and toxin reactivation rates by fecal lysates and to correlate these values with meta-proteomic GUS abundances.^21, 23^ Here we found MPA reactivation rates between 3 nM/sec and 114 nM/sec for MMF-treated transplant recipients’ samples, and between 6 and 18 nM/sec for the healthy individual samples (Figure 4B). Thus, a range of values were observed across all samples, and the highest rates recorded were from MMF-treated transplant recipient samples. However, the > 20-fold differences in MPA reactivation rates between samples did not correlate with abundance of microbial class, phylum, family, genus, or species (Supplemental Table 4), or with any feature of the metagenomic GUSome profiles outlined in Figure 3 (Supplemental Figure 10), with one exception at the species level. The relative abundance of *Streptococcus parasanguinis*, a species only present in transplant recipients, correlated with rate of MPA reactivation with a slope that was significantly non-zero (P=0.0486; Supplemental Table 4). However, the relative abundance of this species ranged from just 1-7%, and GUS genes from this species were not detected. Thus, metagenomic sequencing data, even when examined at the level of GUS protein functional classes, are insufficient to explain the differences in MPA reactivation rates observed within or between transplant recipient and control samples. This observation suggests that differences in the expression or abundance of specific gut microbial GUS proteins between samples may provide this explanation.

**Figure 4.**
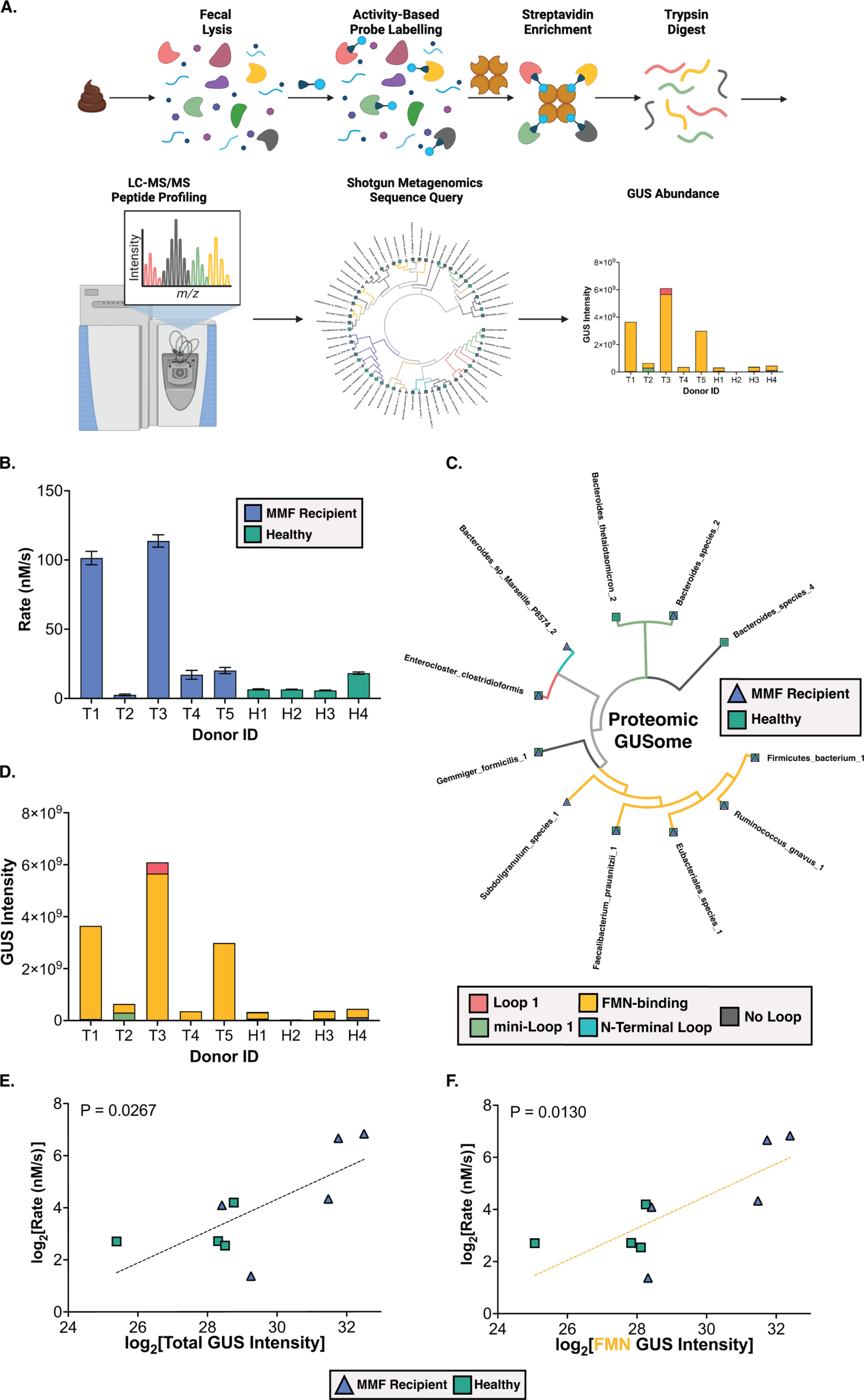
(A) Activity-based probe-enabled proteomic pipeline. (B) *Ex vivo* reactivation of MPAG by donor fecal extracts; data reflects the mean of three biological replicates and error bars reflect SEM. (C) Cladogram reflecting GUS proteins identified across cohort. GUS class and treatment groups from which proteins were derived are indicated. (D) Proteomic profiles for GUS proteins in MMF recipients and healthy individuals. Metaproteomic GUS abundance is represented by Intensity, which is the combined peptide signal intensity corresponding to each reference GUS sequence from Shotgun Metagenomic Sequencing. Proteins were binned according to GUS class. (E-F) Correlation analysis between MPAG reactivation rate and normalized total GUS protein abundance (E; P = 0.027) or FMN GUS protein abundance (F; P = 0.013). P values reflect confidence in a slope that is significantly non-zero. Figure 4A was created with BioRender.com.

### MMF Treatment and Gut Bacterial GUS Protein Composition

Next, we sought to explore the metaproteome of GUS enzymes between the fecal samples collected from the five kidney transplant recipients receiving MMF and from the healthy individuals. The protein lysates from each fecal sample were examined by activity-based probe-enabled proteomics using a biotin-linked covalent probe for GUS enzymes, which we have adapted from our previous studies using human fecal samples and therapeutic reactivation rates, and the reactivation of the antimicrobial compound triclosan.^21, 23^ The proteomics pipeline outlined in Figure 4A was followed to output peptide fragments that were then used to identify individual GUS proteins.^21, 23^ Importantly, though, the current study uses the protein sequences derived from shotgun metagenomics collected from these exact fecal samples (Figures 2 and 3) as the reference database to identify GUS proteins, rather than a standard reference metagenome like the Integrated Gene Catalog (IGC).^28^ In doing so, we found that GUS proteome percent peptide coverage was significantly improved as determined by a by a Wilcoxon matched-pairs signed rank test (P = 0.031; Supplemental Figure 11) using the cohort-specific metagenomics data compared to the IGC as a reference database.

GUS proteins of all structural classes that were detected in metagenomics were also present in the probe-enabled proteomics data, except for Loop 2 and mini-Loop 2 (Figure 4C). Eleven unique GUS proteins were detected in total, with most GUS proteins being present in both groups (Figure 4C). All samples contained GUS proteins, with FMN-binding GUS enzymes being detected the most frequently across the cohort (Figure 4D). Notably, samples from transplant recipients receiving MMF contained significantly more GUS proteins than healthy individuals when compared by Welch’s t-test (P = 0.034; Supplemental Figure 12). We then compared abundance of GUS proteins by structural class and found that FMN-binding GUS enzymes are significantly elevated in transplant recipients receiving MMF compared to healthy individuals as determined by a Welch’s t test (P = 0.029; Supplemental Figure 12). In addition, “No Loop” GUS enzymes were abundant in the four healthy individual samples but were not detected in four of the five MMF recipients (P = 0.009; Supplemental Figure 12). Outside of FMN-binding and “No Loop” GUS enzymes, there were no other significant differences in GUS abundances by structural class between treatment groups (Supplemental Figure 12). Thus, activity-based probe-enabled proteomics demonstrate that the fecal samples from transplant recipients who received MMF showed increased levels of FMN-binding GUS enzymes compared to fecal samples from healthy individuals.

To directly evaluate the impact that these differences in GUS abundance might have on MPAG processing in the gut, we sought identify trends between MPA reactivation rates and GUS composition and/or abundance by sample. First, we found that reactivation linearly increases with overall abundance of GUS proteins regardless of treatment group with a correlation that is significantly non-zero (p = 0.027; Figure 4E). We then mapped abundance of GUS proteins by structural class to rate of MPA reactivation. Strikingly, the abundance of FMN-binding GUS enzymes also strongly correlates with rate of MPAG reactivation with a slope that is significantly non-zero (p = 0.013; Figure 4F). No significant correlations were observed between proteomic levels of other GUS structural classes and MPA reactivation (Supplemental Figure 13). Together, these results indicate that FMN-binding GUS enzymes are associated with reactivation of MPA in human fecal samples.

### FMN-Binding GUS Active Site Environment and MPAG

To identify a structural rationale for the strong positive correlation of abundance of FMN-binding GUS enzymes with MPA reactivation rate, we used AlphaFold to model protein structures for the five FMN-binding GUS enzymes identified by the activity-based probe-enabled proteomics pipeline employed here.^29^ We overlaid these five structures with extant structures of FMN-binding GUS enzymes from *Roseburia hominis* 2 GUS and *Faecalibacterium prausnitzii* L2-6 GUS, which were resolved with x-ray crystallography (Figure 5A).^26^ All core protein domains were heavily conserved between the crystal structures of FMN GUS enzymes and our AlphaFold models, which is reflected in the RMSD values ranging from 0.5 Å to 2.3 Å (Figure 5B). The C-terminal domain (CTD) flanking the enzyme active site is the area with the greatest variance in positioning between our GUS enzyme models, which is expected as this domain has yet to be resolved in a crystal structure of an FMN-binding GUS. Overall, the models are similar, as reflected in the relatively low RMSDs and high percent identities for these FMN-binding GUS enzymes identified in the metaproteome (Figure 5B and 5C).

**Figure 5.**
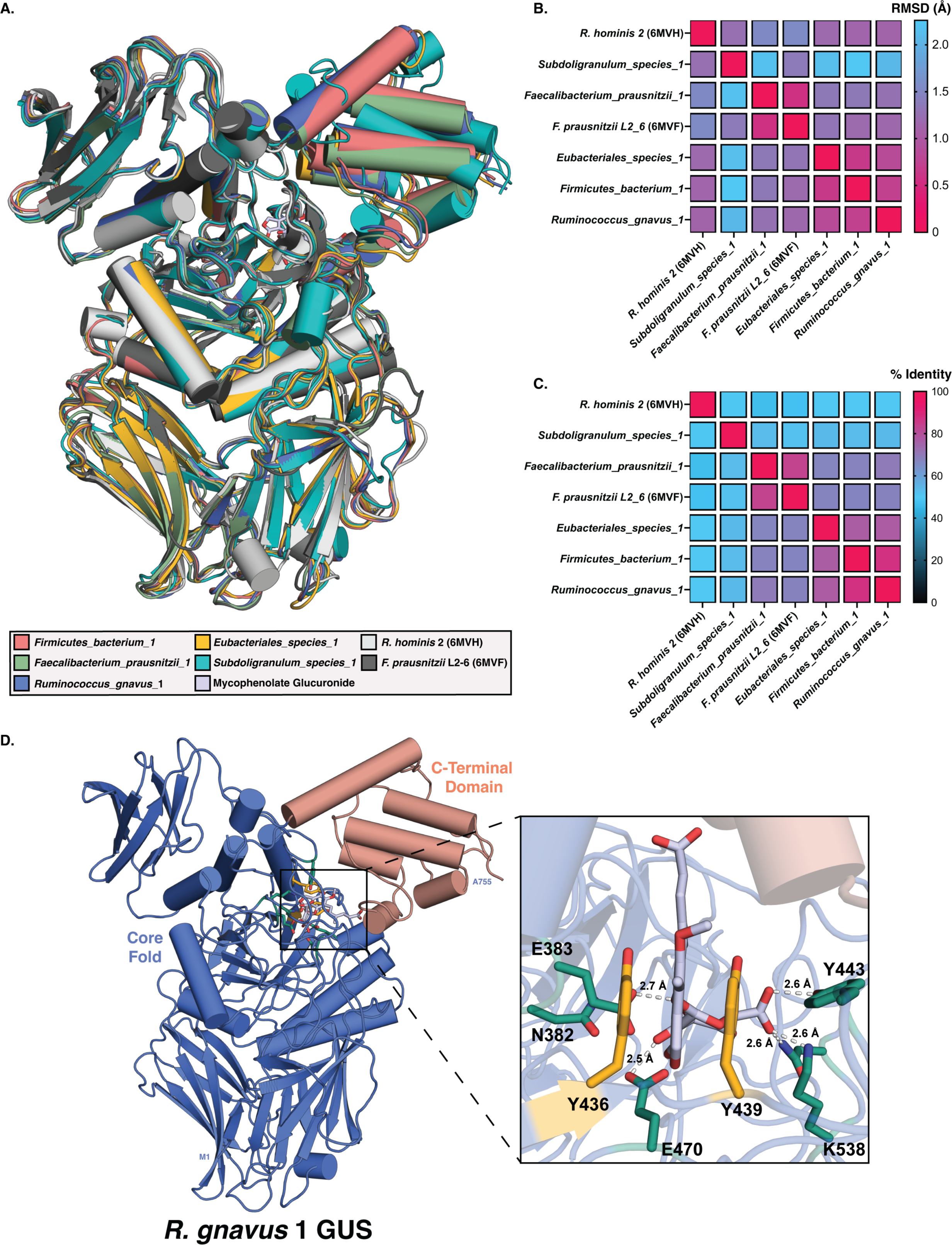
Comparison of AlphaFold models for five meta-proteome derived FMN-binding GUS enzymes with x-ray crystal structure models for two FMN-binding GUS enzymes. (A) Overlay of seven structures. (B) Heatmap comparing Root-mean-square deviation (RMSD) in Angstroms for the seven structures. (C) Heatmap comparing percent sequence identity for the sequences of the seven structures. (D) Active site analysis of R. gnavus 1 GUS monomer (blue) with C-terminal domain shown in coral. Catalytic residues are shown in green, conserved aromatic residues are shown in yellow, and MPAG is shown in gray.

We next analyzed the active sites of our modelled FMN-binding GUS enzymes to identify residues that may be involved in the efficient hydrolysis of MPAG. MPAG was docked into the active sites of each of the modeled structures for the proteome-derived FMN-binding GUS enzymes. Intermolecular interactions predicted between MPAG and *R. gnavus* 1 GUS are shown in Figure 5D, which are representative of the interactions predicted between MPAG and all the FMN-binding GUS enzymes examined. Two consistent interactions were then noted across the substrate-enzyme complexes. First, the MPA core scaffold is positioned between two aromatic residues favorable for π-π stacking interactions. Importantly, while tyrosine 439 is conserved across all GUS enzymes, the aromatic moiety of tyrosine 436 is only conserved across FMN-binding GUS enzymes and not across GUS enzymes from other structural classes. Second, the carboxylic acid tail of MPA is positioned approximately 3 Å from the CTD (shown in coral in Figure 5D), providing conditions favorable for salt bridges or Van Der Waals interactions between the carboxylic acid and residues of the CTD. Together, these structural models indicate that aromatic residues conserved in the active sites of FMN-binding GUS enzymes provide χ-χ stacking interactions that may efficiently facilitate MPA reactivation by these gut microbial proteins.

## Discussion

To identify the GUS genes within our shotgun metagenomics data, we applied a structure-guided approach towards our cohort’s predicted protein sequences (Figure 3A).^24, 26^ We identified 64 unique GUS genes in total from 7 distinct structural classes, with the majority being derived from *Bacteroidetes* and *Firmicutes*. Several clusters of GUS genes of the same structural class were notably unique or absent in transplant recipients altogether (Figure 3B). We sought, then, to explain the range in MPA reactivation by fecal lysates using either overall microbiome composition or by metagenomic GUS profiles between samples or groups, as metagenomic gene abundances have historically been used to predict gene expression and functional potential.^8, 30–33^ However, we were unable to find any correlations between the rates of drug reactivation measured and any compositional or GUS metagenomic feature (Supplemental Figure 10). We therefore concluded that, for this cohort, the fecal shotgun metagenomic data were insufficient to pinpoint specific gut microbial GUS enzymes that may have been responsible for the 38-fold difference in MPAG to MPA processing rates observed between fecal samples.

We then used activity-based probe-enabled proteomics to directly identify and quantify the GUS proteins present in our cohort samples (Figure 4A).^34^ Peptide fragments were matched to eleven unique GUS proteins, with most structural classes of GUS proteins being detected in both treatment groups, and FMN-binding GUS enzymes being the most prevalent (Figure 4C). Samples from transplant recipients receiving MMF contained more GUS proteins overall than healthy individuals (P=0.034; Supplemental Figure 12). Furthermore, FMN-binding GUS enzymes were more abundant in MMF transplant recipient samples (P=0.029), and “No Loop” GUS enzymes were not detected in the majority of MMF recipients (P=0.0097). When these findings are considered in the context of our metagenomics data, two aspects are noteworthy. First, while the metagenomics showed that samples from all members of the cohort except T3 contained at least one “No Loop” GUS gene, proteins from this structural class were only detected in one MMF recipient by metaproteomics. Conversely, “No Loop” GUS proteins were detected in both the metagenomics and the metaproteomics of all healthy individuals (Figure 3D; Figure 4D). In all previous studies quantifying GUS abundance with targeted metaproteomics, “No Loop” GUS enzymes were detected in all samples.^21, 34^ Therefore, the intestinal environment of transplant recipients receiving MMF appears to shift in favor of FMN-binding GUS enzymes over “No Loop” GUS ortholog. Second, the eleven unique GUS proteins were identified using metaproteomics represents a small subset of the 64 unique GUS genes that could be expressed across the cohort. Our previous work has shown that all structural classes of GUS enzymes are detectable by our probe, so the differences are not driven by differential probe reactivity. Instead, our results suggest that only this subset of GUS enzymes were expressed with sufficient abundance to be detected in our probe-enabled proteomics pipeline (Figure 4A).^23^ By using the protein sequences derived from shotgun metagenomics collected from these fecal samples as the reference database to identify GUS enzymes (Figures 2 and 3), we significantly improved peptide coverage for matched proteins. Thus, while fewer unique proteins are abundant than are encoded within the metagenomic GUSome for our cohort (Figure 3B), the metaproteomics peptide coverage suggests high confidence matches for these eleven proteins.

When rates of MPA reactivation were compared to these abundances, we found a positive correlation between rate and overall abundance of GUS (Figure 4E). By further delineating this correlation according to individual structural classes of GUS, we identified FMN-binding GUS enzymes as the driving force behind this positive correlation. Indeed, when rates of MPA reactivation are compared to FMN-binding GUS abundance, we see the strongest positive correlation between abundance and rate of MPA reactivation for any class of GUS (Figure 4F; Supplemental Figure 13). We attempted to relate the relative gene abundance for any structural class of GUS as determined by metagenomics with the protein abundance of the class as determined by metaproteomics, but no correlations were identified (Supplemental Figure 14). We extended our approach towards relating bacterial abundance at any taxonomic level with rates of MPA reactivation; again, though, no significant trends were present for bacteria containing GUS genes (Supplemental Table 4). Together, these results suggest that changes in FMN-binding GUS protein expression, but not the overall abundance of these GUS-expressing microbes, are responsible for reactivation of MPA. Indeed, treatment with MMF has been shown to broadly alter the gene expression profiles of bacteria within the gut.^35^ While metagenomic approaches provide an accurate map of the possible proteins within one’s microbiota and are uniquely valuable in their ability to improve specificity of metaproteomic peptide fragment mapping, neither the relative abundance of bacteria or the abundance of genes should be used as a metric to predict GUS protein expression or activity.

Our study is the first to apply GUS-targeted metaproteomics towards analyzing a cohort who routinely use MMF. By doing so, we highlighted stark differences between the GUS profiles of these groups, suggesting that GUS abundance is increased in renal transplant recipients, and that FMN-binding GUS enzymes are the driving force behind the reactivation of MPA. All members of the transplant recipient cohort contain at least one FMN-binding GUS in their metaproteomics data, and the rate of MPA reactivation increases with abundance of FMN-binding GUS enzymes. However, samples from healthy individuals have a lower abundance of FMN-binding GUS enzymes and generally reactivate MPA less efficiently compared to the samples from MMF recipients. In all prior studies that apply activity-based proteomics towards discovering structural classes of GUS responsible for drug reactivation, “Loop 1” GUS enzymes were most strongly correlated with increased rate of reactivation.^21, 34^ Some classes of GUS enzymes were sparsely represented across our cohort as a whole (Figure 4; Supplemental Figure 11), with no “Loop 2” GUS enzymes being detected and with “Loop 1” GUS enzymes only being detected in four of nine samples. The increased abundance of FMN-binding GUS enzymes in MMF recipients relative to healthy individuals supports our conclusion that FMN-binding GUS promote MPA reactivation in the gut, which may in turn contribute to GI toxicity and systemic recirculation of active MPA.

Finally, to pursue a structural rationale for the strong positive correlation of abundance of FMN-binding GUS with MPA reactivation, we modeled the proteomic GUSome FMN-binding GUS enzymes using AlphaFold, then overlaid the structures with extant structures of FMN-binding GUS enzymes that were resolved with x-ray crystallography (Figure 5A).^29^ Each of our models position the flexible CTD directly outside of the active site, where it may act as a “gate” for glucuronidated substrates. Given that removal of the CTD diminishes all GUS activity, these initial interactions between the CTD and substrate are likely integral to substrate recruitment and positioning for hydrolysis.^20, 21, 23^ Within the active site, and where the accuracy of the AlphaFold models is expected to be the highest, MPAG likely interacts via ρ-ρ stacking with two aromatic residues residing above the back of the glucuronic acid catalytic site (yellow; Figure 5D). While one of these aromatic residues (Y439 in *R. gnavus* 1 GUS) is conserved across all GUS enzymes, this second aromatic residue is notably not conserved in the “No Loop” GUS enzymes that are closest to FMN-binding GUS proteins by sequence identity. The positioning of these aromatic residues is similarly reflected in the extant structures of FMN-binding GUS enzymes, corroborating the accuracy of our AlphaFold models in this region. Together, these observations suggest that MPAG is efficiently reactivated by FMN-binding GUS enzymes due to conditions favorable for ρ-ρ stacking at the active site. Future studies will further explore the molecular rationale of MPA reactivation by FMN-binding GUS enzymes.

MMF is an important immunosuppressive agent administered to organ transplant recipients and is widely prescribed to treat autoimmune disorders, with many transplant recipients requiring drug administration long-term. Here, we show that fecal samples with a greater abundance of FMN-binding GUS enzymes, particularly those from MMF-treated transplant recipients, reflected a faster rate of reactivation of MPA compared to healthy individuals. Together, our findings demonstrate that gut microbial FMN-binding GUS enzymes efficiently reactivate MPA, which may play a significant role in MPA-induced GI toxicity. The data presented in this study reinforce the relevance of the microbiome in MPA-induced toxicity. However, there are several limitations to this study. First, the sample size is small, which was by design to validate our proteomics pipeline with an initial set of patient and non-patient samples. A larger cohort of samples will be required to further validate our findings. Second, the patient samples were collected in New York while the non-patient samples were collected in North Carolina, and difference in geography can affect human gut microbiomes.^36–39^ Third, the patients were suffering from kidney failure prior to transplant, and then had in most cases just undergone the transplant surgery. Both factors are likely to influence the structure and activity of the gut microbiome, as will their associated use of therapeutics beyond mycophenolate. Fourth, the fecal samples were not handled after collection to allow for transcriptomics analysis toward determining whether transcript levels might correlate with fecal MPA reactivation rates. Finally, all subjects were male to match MMF-recipient with MMF-recipient characteristics but leaving sex as a biological variable unexamined. Despite these limitations, several clear conclusions can be drawn from the data collected, most notably that the proteomics provided correlations with drug reactivation that the metagenomics did not. Thus, with additional validation and development, the data presented here may lay the groundwork for the identification of transplant recipients at-risk for MPA-induced gut damage or, by extending the established concept of targeted microbial GUS inhibitors, towards alleviating the GI toxicity in patients receiving MMF therapy.

### Method Details

#### Fecal sample collection

From May 2019 to January 2020, five male kidney transplant recipients were enrolled for collection of fecal specimens. The Weill Cornell Institutional Review Board approved the study (IRB#1207012730) and all transplant recipients provided written informed consent. Fecal samples were also collected from four healthy male volunteers at the University of North Carolina at Chapel Hill (IRB#17-1528). Fecal samples were collected using a toilet specimen collection kit (Fisher Scientific) and were stored at -80 °C until further use. Demographics of the cohort can be found in Supplemental Table 1.

#### Preliminary Metagenomics Analysis

Metagenomics samples were processed through the pipeline shown in Supplemental Figure 1. Raw metagenomics genes were trimmed, filtered, and annotated, then assembled into gene and protein sequences using Metagenomics Analysis Toolkit (MOCAT2 v2.0.1).^40^ To determine the relative abundance of bacterial taxa for each sample, paired-end reads were analyzed using Metaphlan (v3.0.13) and results were graphed using ggplot2 (v3.3.5) in R (v4.1.2), as shown in Figure 2 and Supplemental Figures 2 – 5.^41, 42^ MetaPhlan (v3.0.13) was also used to generate a Biological Observation Matrix (BIOM) for each sample, which was processed with QIIME2 (v2022.2.0) for diversity analyses.^41, 43^ Alpha diversity was determined using the “diversity alpha” function and the Shannon Diversity Index parameter in QIIME2.^43^ Beta diversity was calculated using Bray-Curtis equilibrium distances and plotted with the Constrained Analysis of Principal Coordinates ordination method using the “capscale” function within the Vegan (v2.5-7) package in R (v4.1.3), as described previously.^44^ The Bray-Curtis distances were compared via PERMANOVA to assess differences in species-level composition between kidney transplant recipients and healthy individuals.^44^

#### Metagenomics Gene Abundance

Nucleotide sequences were used to generate gene indices and Sequence Alignment Maps (SAM) using Bowtie2 (v2.4.1).^45^ Amino acid and nucleotide sequences were used to generate Generic Feature Format (GFF) files using PRODIGAL (v2.6.3).^46^ The paired genes from the SAMs were assigned to genes within the GFF files and counted using the SUBREAD package featureCounts (v2.0.0).^47^ Read counts were converted to relative counts based on the total count scaling factor to account for differences in read numbers between samples, using the formula below as previously described.^24, 48^ Relative count was determined as follows:

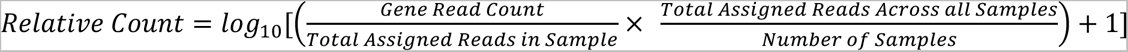

To assess gene length bias in read counts, relative counts were plotted as a function of gene length. To negate the bias, we further scaled the relative counts using the slope of the gene abundance linear regression as follows to reach the final Normalized Gene Abundance:

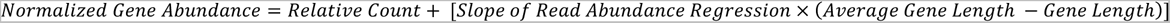

#### Identification and Characterization of GUS Sequences

Metagenomic amino acid sequences were each aligned pairwise to 17 representative GUS enzymes with reported crystal structures using Protein-Protein BLAST (BLASTP v2.5.0+).^49^ Candidate sequences with ≥ 25 % identity to any representative GUS enzyme were then assessed for the presence of 7 conserved residues.^24^ Sequences that both met the identity threshold and contained all 7 conserved residues were accepted as GUS enzymes. Accepted sequences were filtered for redundancies at a sequence identity threshold of 90% using CD-HIT (v4.8.1), and the output was used to form a representative set of GUS sequences for downstream analysis.^50^ Accepted sequences were aligned to representative sequences from each loop class in a Multiple Sequence Alignment (MSA), and GUS class was assigned according to parameters reported previously.^24–26^ Taxonomy was assigned to representative GUS sequences using BLASTP (v2.5.0+) as reported previously, and taxonomic identifiers were used to rename these sequences.^24, 49^ Representative sequences were clustered using the EMBL-EBI search, which was combined with the GUS class and taxonomy to create cladograms using ggtree (v3.2.1) and ggplot2 (v3.3.5) in R (v4.1.2).^42, 51, 52^ Normalized gene abundances for GUS sequences were mapped to their corresponding GUS class then summed to form the plots in Figure 3C and Supplemental Figure 7. All relevant data including gene sequences, loop class, detailed taxonomy, and final gene counts can be found in Supplemental Table 2. Gene_ID’s can be matched to their corresponding “Manuscript ID” using the “Gene_Barcode_Identifiers” tab.

#### Complex Protein Lysate Preparation

Human fecal samples were processed as previously described (21). 5 - 10 g of thawed fecal material collected from each donor was resuspended in 25 mL cold extraction buffer (25 mM HEPES pH 6.5, 25 mM NaCl, one Roche Complete EDTA-free protease inhibitor tablet in 50 mL buffer) and 500 mg autoclaved garnet beads then vortexed. Samples were centrifuged at 300 x *g* for 5 mins at 4 °C and supernatant was collected. 25 mL cold extraction buffer was added to the centrifuged pellet, which was again vortexed and centrifuged. Both supernatants were combined and centrifuged at 300 x *g* for 5 mins at 4 °C two additional times to further remove insoluble fiber. The supernatant was then sonicated twice on a Fischer Scientific Sonic Dismembrator Model 500 with 0.5 second pulses for 1.5 mins and the lysate was mixed by inversion between each sonication. Lysate was then centrifuged at 17,000 x *g* for 20 mins at 4 °C to remove insoluble debris then decanted. The lysate was then concentrated with Amicon Ultra 15 mL 30 kDa centrifugal filters and exchanged with fresh extraction buffer three times to remove metabolites. After buffer exchanging, the total protein concentration of the final fecal lysate for each sample was measured with a Bradford assay using purified *Escherichia coli* β-Glucuronidase as a nitrogen and stored at -80 °C until later use in proteomics and fecal lysate assays.

#### GUS activity-based probe (ABP)

Cyclophellitol-based probe JJB397 was synthesized and purified as previously described to form a biotin-linked covalent inhibitor of GUS enzymes.^27^

#### Metaproteomics

General Proteomics workflow is shown in Figure 4A and was adapted from our previously reported GUS-targeted Activity Based Proteomic Profiling pipeline.^34^ Human fecal extracts (3 mg total protein) were thawed then incubated at 37 °C for 60 mins with 10 μM biotin-linked ABP JJB397 in 500 μL cold extraction buffer (25 mM HEPES pH 6.5, 25 mM NaCl, 1% DMSO final, one Roche Complete EDTA-free protease inhibitor tablet in 50 mL buffer). Reactions were quenched by adding 125 μL Triton + Urea buffer (50 mM HEPES pH 6.5, 125 mM NaCl, 1 mM EDTA, 1 mM EGTA, 1% v/v Triton X-100, 0.1% w/v SDS, DMSO final, 1:500 Roche Complete EDTA-free protease inhibitor tablet, 2M Urea) and heating at 95 °C for 5 min. Samples were cooled on ice then washed five times with Triton + Urea buffer using 0.5 mL Amicon Ultra 10K membrane filters to remove unreacted probe. Between each wash, samples were centrifuged at 13,000 x *g* for 5 mins. The volume of each sample was then adjusted to 1 mL using Triton + Urea buffer. 20 μL per sample MyOne^TM^ Streptavidin T1 (Invitrogen) beads were washed three times in Triton + Urea buffer then diluted to a final volume allowing for 100 μL to be added to each sample. 100 μL of the Streptavidin T1 bead slurry was added to each 1 mL sample and the samples were then rotated end-over-end for 120 mins at room temperature. Beads were then washed five μL NH_4_HCO_3_ (pH 7.8). Between each wash step, streptavidin beads were isolated using a DynaMag^TM^-2 Magnet (Invitrogen). After the final wash step, beads were resuspended in 100 μL NH_4_HCO_3_ (pH 7.8) and stored at -20 °C for proteomic analysis. Samples were subjected to on-bead trypsin digestion, as previously described.^53^ After the last wash buffer step, 50 µl of 50 mM ammonium bicarbonate (pH 8) containing 1 µg trypsin (Promega) was added to beads overnight at 37°C with shaking. The next day, 500 ng of trypsin was added then incubated for an additional 3 h at 37°C with shaking. Supernatants from pelleted beads were transferred, then beads were washed twice with 50 μl LC/MS grade water. These rinses were combined with original supernatant, then acidified to 2% formic acid. Peptides were desalted with peptide desalting spin columns (Thermo) and dried via vacuum centrifugation. Peptide samples were stored at -80°C until further analysis.

#### Metaproteomics LC-MS/MS Analysis

The peptide samples analyzed were by LC-MS/MS using an Easy nLC 1200 coupled to a QExactive HF mass spectrometer (Thermo Scientific). Samples were injected onto an Easy Spray PepMap C18 column (75 μm id × 25 cm, 2 μm particle size) (Thermo Scientific) and separated over a 90 min method. The gradient for separation consisted of 5–40% mobile phase B at a 250 nl/min flow rate, where mobile phase A was 0.1% formic acid in water and mobile phase B consisted of 0.1% formic acid in 80% ACN. The QExactive HF was operated in data-dependent mode where the 15 most intense precursors were selected for subsequent fragmentation. Resolution for the precursor scan (m/z 350–1600) was set to 60,000 with a target value of 3 × 106 ions, 100 ms max injection time. MS/MS scans resolution was set to 15,000 with a target value of Dynamic exclusion was set to 30 s, peptide match was set to preferred, and precursors with unknown charge or a charge state of 1 and ≥ 7 were excluded.

#### Metaproteomics Data Analysis

Raw data were processed as described previously, with the following modifications.^21, 23^ MaxQuant (v1.6.3.4) was used for peptide identification and quantitation.^54^ Data were searched against a reference database containing non-redundant protein sequences derived from metagenomic shotgun sequencing of all samples within the cohort (containing 645,141 entries), the Uniprot Human database (containing 26,122 entries), and a potential contaminants database.^55^ Proteins were filtered for a false discovery rate (FDR) of 1% at the unique peptide level, and potential contaminants and decoys were removed. Peptide peak areas were extracted and summed for each protein and the protein intensities were used for relative quantitation. Proteomic data can be found in Supplemental Table 3. Percent peptide coverage was compared for GUS enzymes identified using either the IGC or cohort metagenomics as the reference database using a Wilcoxon two-tailed t-test; results are shown in Supplemental Figure 11.^28^ Proteomic intensities were log_2_-transformed then compared between groups by Welch’s t-test; results are shown in Supplemental Figure 12. Normalized intensities were compared to normalized gene abundances as determined with metagenomics; P values reflect confidence in a slope that is significantly non-zero and results are shown in Supplemental Figure 4. Normalized intensities were also correlated with normalized rate of MMF processing, with P values reflecting confidence in a slope that is significantly non-zero; results are shown in Figure 4E, Figure 4F, and Supplemental Figure 13.

#### Fecal Lysate MPAG activity

Thawed fecal lysate was diluted to 1 mg/mL in extraction buffer. Solid MPAG was purchased (Toronto Research Chemicals, CAT# M831520) and suspended in 100% DMSO at 50 mM and stored at -80 °C until further use. MPAG (50 mM) was diluted in ddH_2_O to a working concentration of 4 mM MPAG. Final reaction conditions were 0.1 mg/mL fecal lysate and 400 µM MPAG in assay buffer (25 mM HEPES pH 6.5, 25 mM NaCl) at a final volume of 50 µL. 4 mM MPAG (5 µL) was added to 40 µL assay buffer and incubated at 37 °C for 5 mins. The reaction was initiated by adding 5 µL of fecal lysate (1 mg/mL) and incubated at 37 °C. The reaction was quenched with an equivalent volume of 25% trichloroacetic acid at designated timepoints. Each sample had five endpoints (including 0 mins) which were either every 15 mins or 30 mins depending on the rate of reaction. Each quenched reaction was transferred to a 1.7 mL microcentrifuge tube then centrifuged at 13,000 rpm for 20 mins. 80 µL of supernatant was transferred to a high-performance liquid chromatography (HPLC) vial and concentration of MPAG at each timepoint was quantified on an Agilent 1260 Infinity II liquid chromatography system. Samples were stored in an autosampler at 8 °C prior to separation on an Agilent InfinityLab Poroshell 120 C18 column (4.6 x 150 mm, 2.7 µm) at 38 °C with a flow rate of 0.9 mL/min and injection volume of 40 µL. The LC solvents were as follows; Solvent A: ddH_2_O with 0.1% formic acid; Solvent B: 98% acetonitrile with 0.1% formic acid. The LC flow gradient was as follows: constant 98% A / 2% B for 0 - 2 mins, linearly ramp to 2% A : 98% B over 2 - 15 mins, constant 2% A : 98% B for 15 – 23 mins, linearly ramp to 98% A : 2% B for 23 - 25 mins. MPAG was monitored at 280 nm with a reference of 360 nm, eluting at 9.6 mins. The area under the curve of MPAG was converted to concentration of MPAG using a standard curve. MPAG concentration as a function of reaction time was fit to a linear regression, with the slope representing reactivation of MPAG in nM/s. The slopes of three biological replicates were averaged to reach final rates in nM/s then log_2_ transformed. Rates were plotted in GraphPad PRISM GraphPad Prism (v9.3.1) with error bars representing standard error, shown in Figure 4B. Rates were then correlated to relative abundance of bacteria at all taxonomic levels, with P value reflecting confidence in a slope that is significantly non-zero (Supplemental Table 4). Rates were correlated to relative abundance of bacteria at all taxonomic levels, with P values reflecting confidence in a slope that is significantly non-zero; results are tabulated in Supplemental Table 4. Rates were also correlated with relative normalized abundance of GUS genes, with P values reflecting confidence in a slope that is significantly non-zero; results are shown in Supplemental Figure 10. Rates were also correlated with normalized abundance of GUS proteins, with P values reflecting confidence in a slope that is significantly non-zero; results are shown in Figure 4E, Figure 4F, and Supplemental Figure 13.

#### Protein Modelling

The complete amino acid sequences for the five FMN-binding GUS enzymes detected using metaproteomics were modelled as monomers using Alphafold (v2.0).^29^ The models were aligned to *Roseburia hominis* 2 GUS (PDB: 6MVH) and *Facaelibacterium prausnitzii* L2-6 GUS (PDB: 6MVG) using the cealign plugin in PyMOL (v2.5.2).^26^ MPAG was manually docked into the active site in PyMOL using the crystal structure of *Eubacterium eligens* beta-glucuronidase bound to glucuronic acid (PDB: 6BJQ) overlaid to the structures of FMN-Binding GUS enzymes (PDB: 6MVH and PDB: 6MVG) as a reference scaffold.^26, 56^

## Data Availability

Metagenomic and metaproteomic sequencing data derived from the kidney transplant patients will
be made available at accession number phs001879 in the database of Genotypes and Phenotypes
(dbGaP) upon publication.57 Approval from a local institutional review board approval will be needed to obtain the data. Sequencing data derived from healthy individuals will be made available at the Integrated Data Management and Comparative Analysis System for Microbial Genomes and Microbiomes Database (IMG/MER) upon publication. The metaproteomics data and protein
sequence database for healthy individuals [will be] deposited to the ProteomeXchange Consortium
via the PRIDE repository.

## Acknowledgements

We would like to acknowledge Dr. Manikkam Suthanthiran for his guidance and for providing the infrastructure for the biobank collection. We also would like to thank the UNC Microbiome Core Facility (a member of UNC Center for Gastrointestinal Biology and Disease and the UNC Nutrition Obesity Research Center) for metagenomic processing and sequencing. This research is based in part upon work conducted using the UNC Proteomics Core Facility, which is supported in part by P30 CA016086 Cancer Center Core Support Grant to the UNC Lineberger Comprehensive Cancer Center. We would also like to thank current and previous Redinbo lab members for helpful discussions. Figures 3A, 4A, and Supplemental Figure 2 were created with BioRender.com.

## Author contributions

J.B.S., M.M.B., and M.R.R. conceptualization; J.B.S., J.J.S., and A.L.G. data curation; J.B.S. formal analysis; J.B.S., J.J.S., M.M.B., R.L.J., and A.L.G. investigation; J.B.S., M.M.B., and M.R.R. writing – original draft; J.B.S., M.M.B., A.L.G.,J.R.L, and M.R.R. writing – review and editing; J.B.S. and M.R.R. supervision; J.B.S. and M.M.B. methodology; J.R.L. and M.R.R. funding acquisition; M.R.R. project administration.

## Availability of Data

Metagenomic and metaproteomic sequencing data derived from the kidney transplant patients will be made available at accession number phs001879 in the database of Genotypes and Phenotypes (dbGaP) upon publication.^57^ Approval from a local institutional review board approval will be needed to obtain the data. Sequencing data derived from healthy individuals will be made available at the Integrated Data Management and Comparative Analysis System for Microbial Genomes and Microbiomes Database (IMG/MER) upon publication.^58^ The metaproteomics data and protein sequence database for healthy individuals [will be] deposited to the ProteomeXchange Consortium via the PRIDE repository.^59^

**Supplemental Figure 1.**
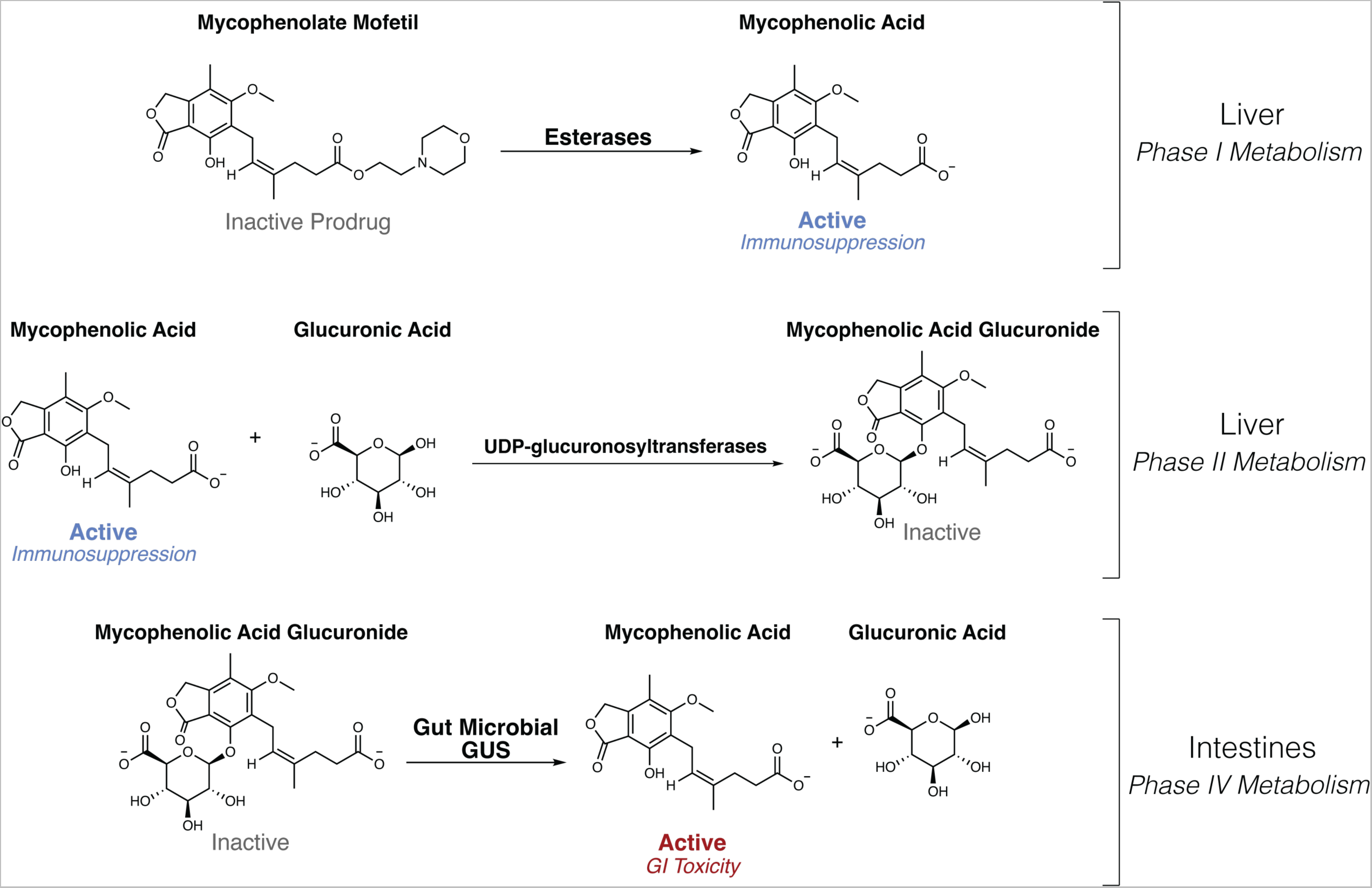
The orally administered MMF is activated by host esterases to mycophenolic acid (MPA), an immunosuppressant that impedes DNA synthesis in B and T lymphocytes. Liver MPA is inactivated via glucuronidation to mycophenolic acid glucuronide (MPAG) by UDP-glucuronosyltransferases (UGTs) and sent to the intestines for excretion. Gut microbial β-glucuronidase (GUS) enzymes remove the glucuronide as a source of carbon, and active MPA is reabsorbed in the gut lumen, contributing to gastrointestinal toxicity.

**Supplemental Figure 2.**
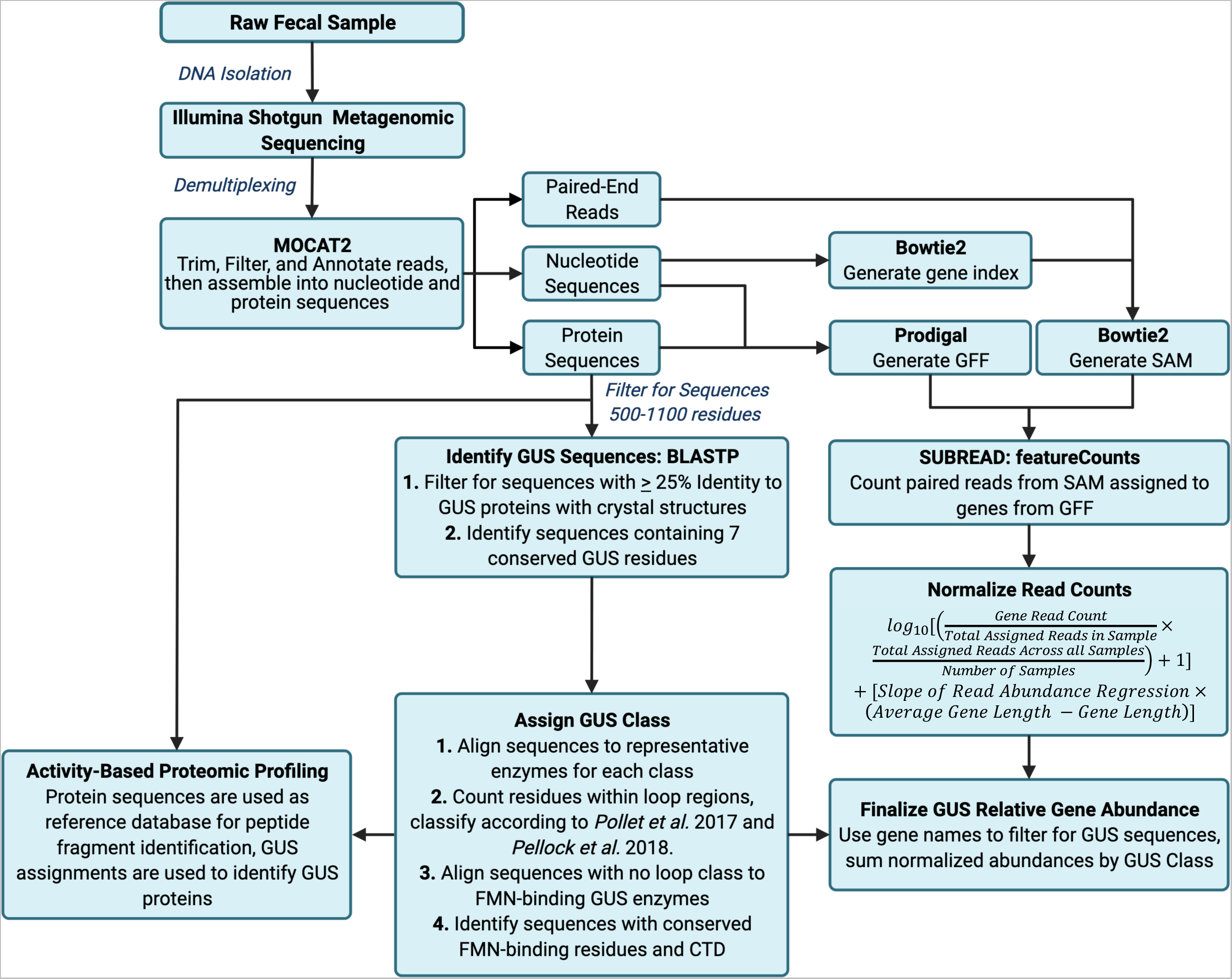
Metagenomic workup and GUS analysis pipeline for human fecal samples processed by Whole Genome Metagenomic Sequencing. Created with BioRender.com.

**Supplemental Figure 3.**
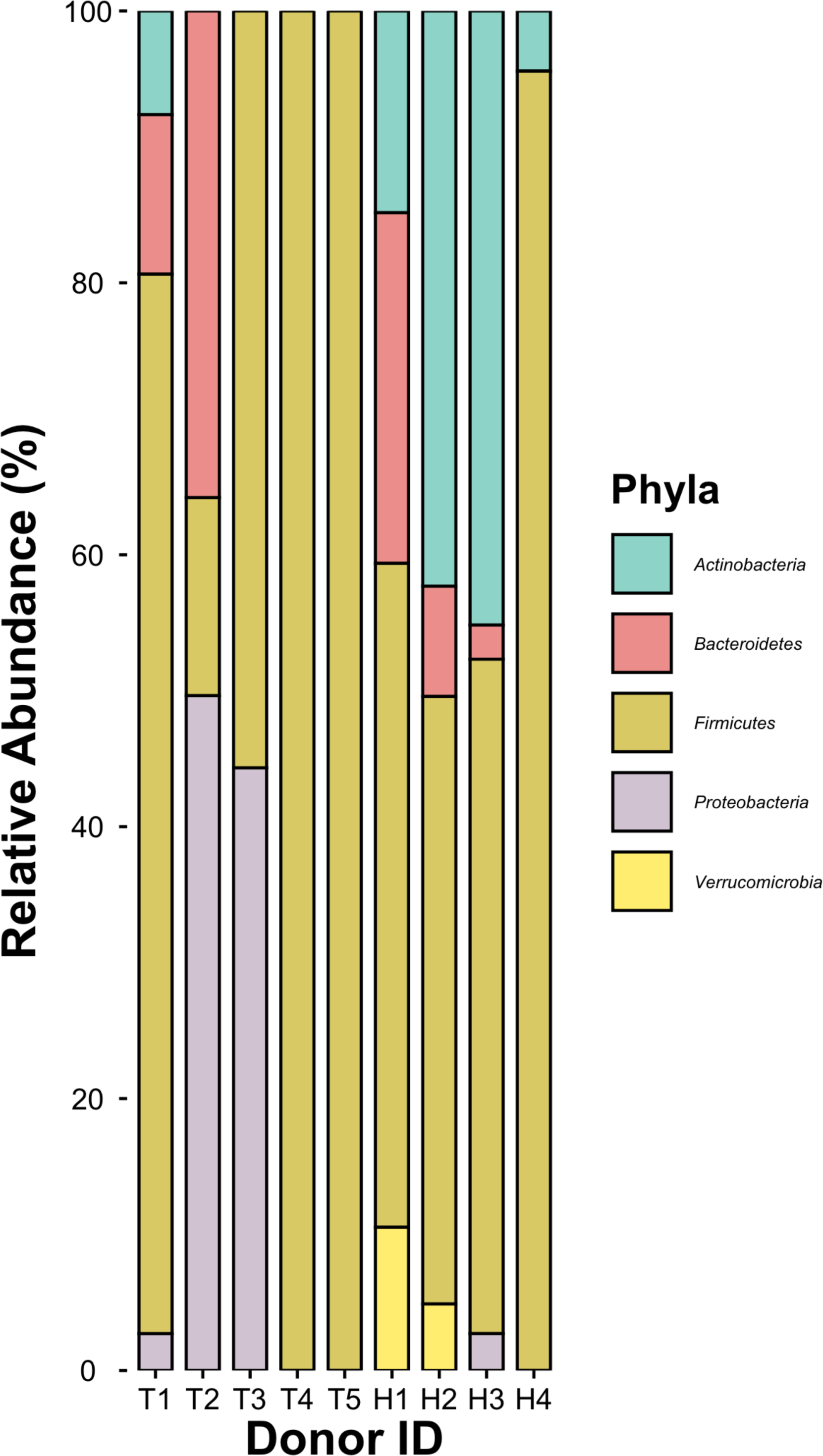
Relative metagenomic abundance of bacteria organized by Phyla. T1-T5 are kidney transplant recipients receiving MMF and H1-H4 are healthy individuals.

**Supplemental Figure 4.**
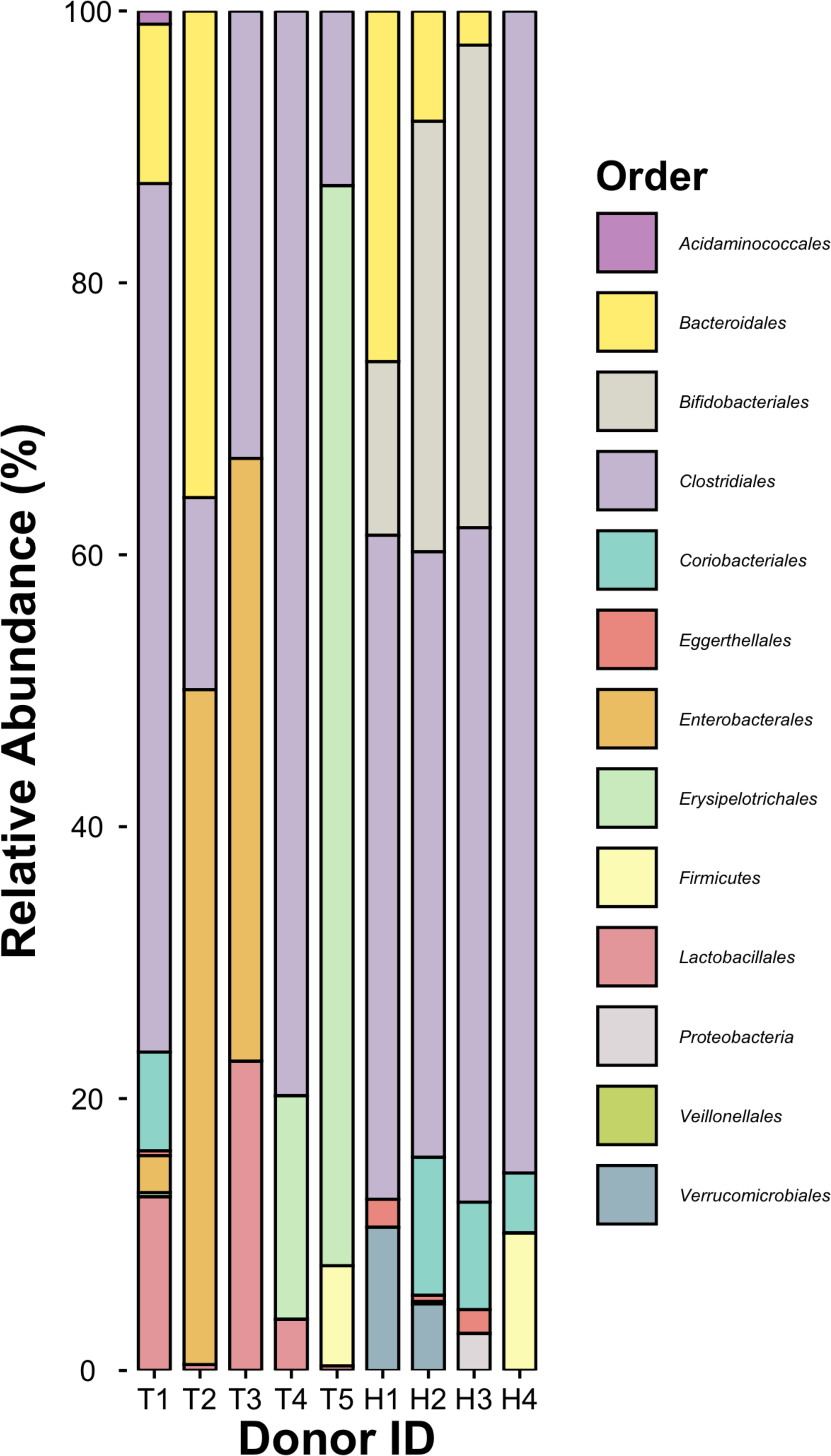
Relative metagenomic abundance of bacteria organized by Order. T1-T5 are kidney transplant recipients receiving MMF and H1-H4 are healthy individuals.

**Supplemental Figure 5.**
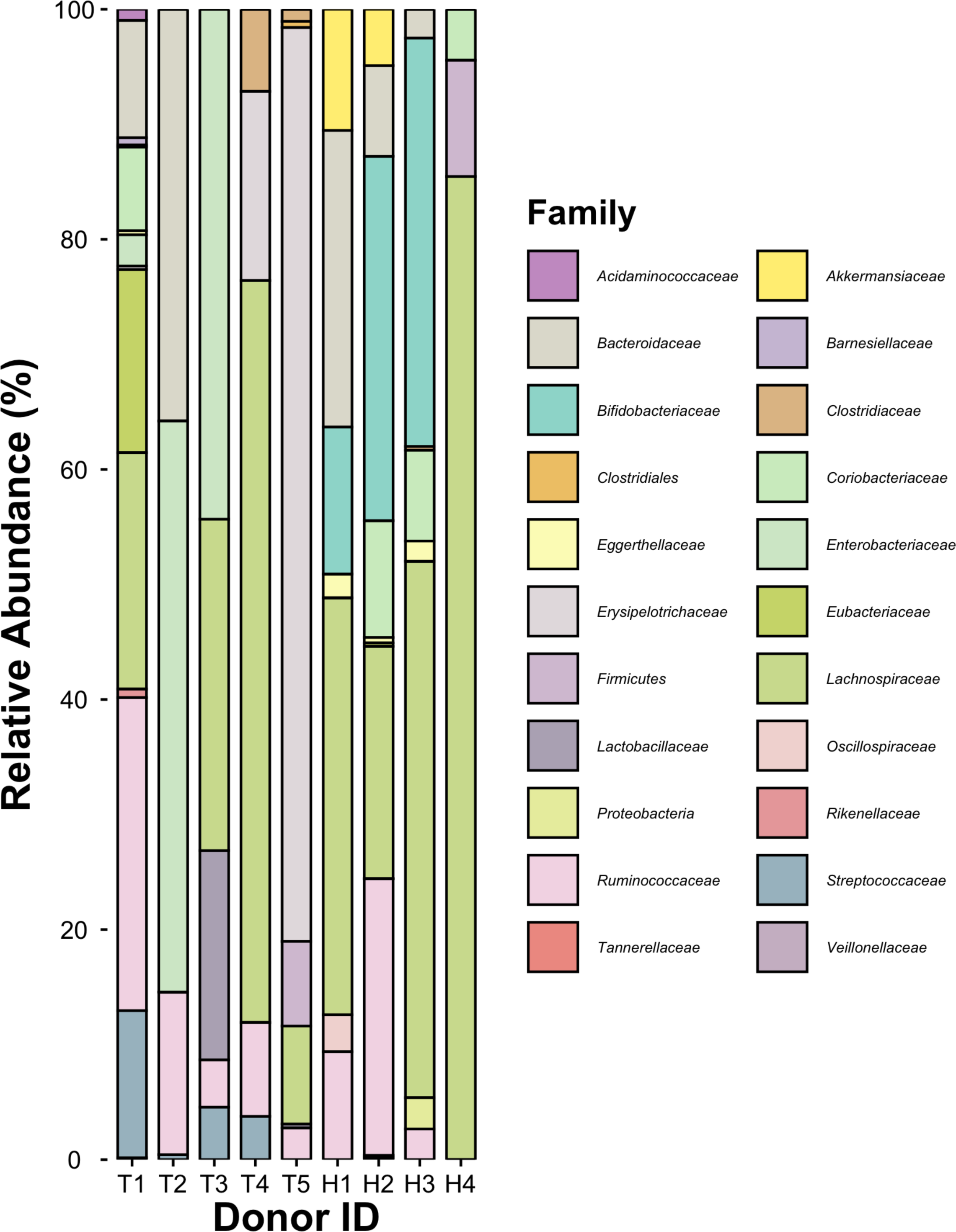
Relative metagenomic abundance of bacteria organized by Family. T1-T5 are kidney transplant recipients receiving MMF and H1-H4 are healthy individuals.

**Supplemental Figure 6.**
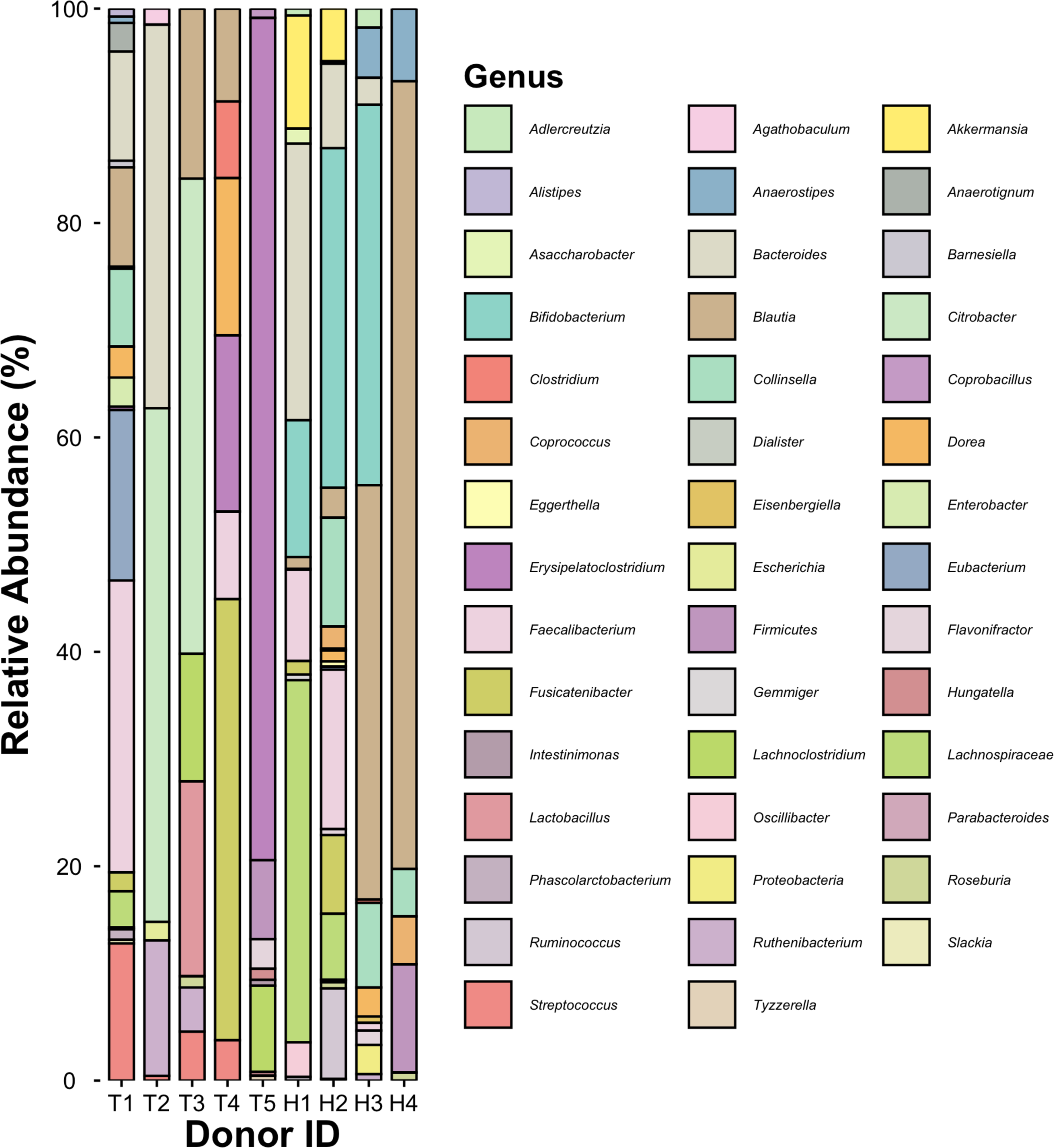
Relative metagenomic abundance of bacteria organized by Genus. T1-T5 are kidney transplant recipients receiving MMF and H1-H4 are healthy individuals.

**Supplemental Figure 7.**
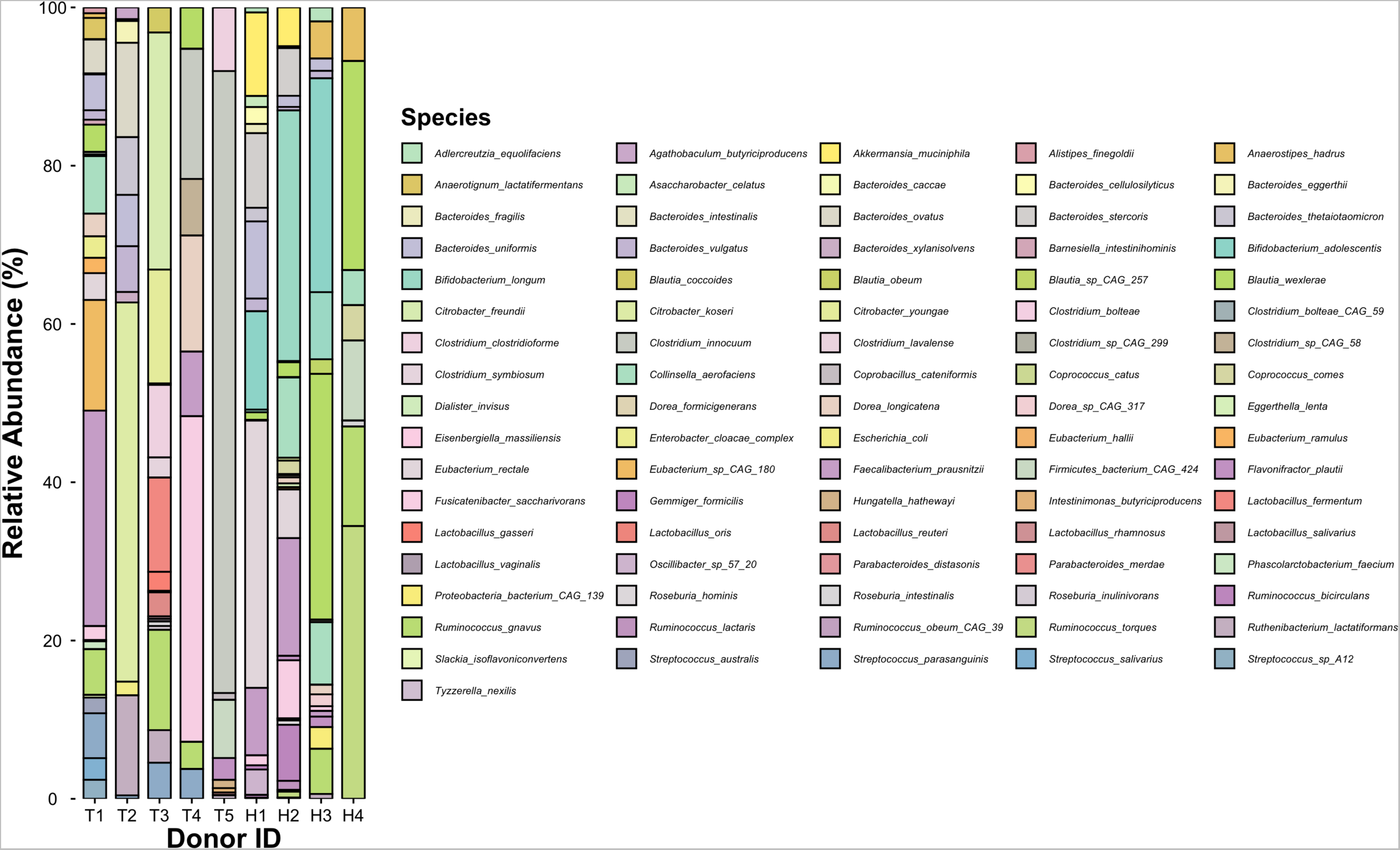
Relative metagenomic abundance of bacteria organized by Species. T1-T5 are kidney transplant recipients receiving MMF and H1-H4 are healthy individuals.

**Supplemental Figure 8.**
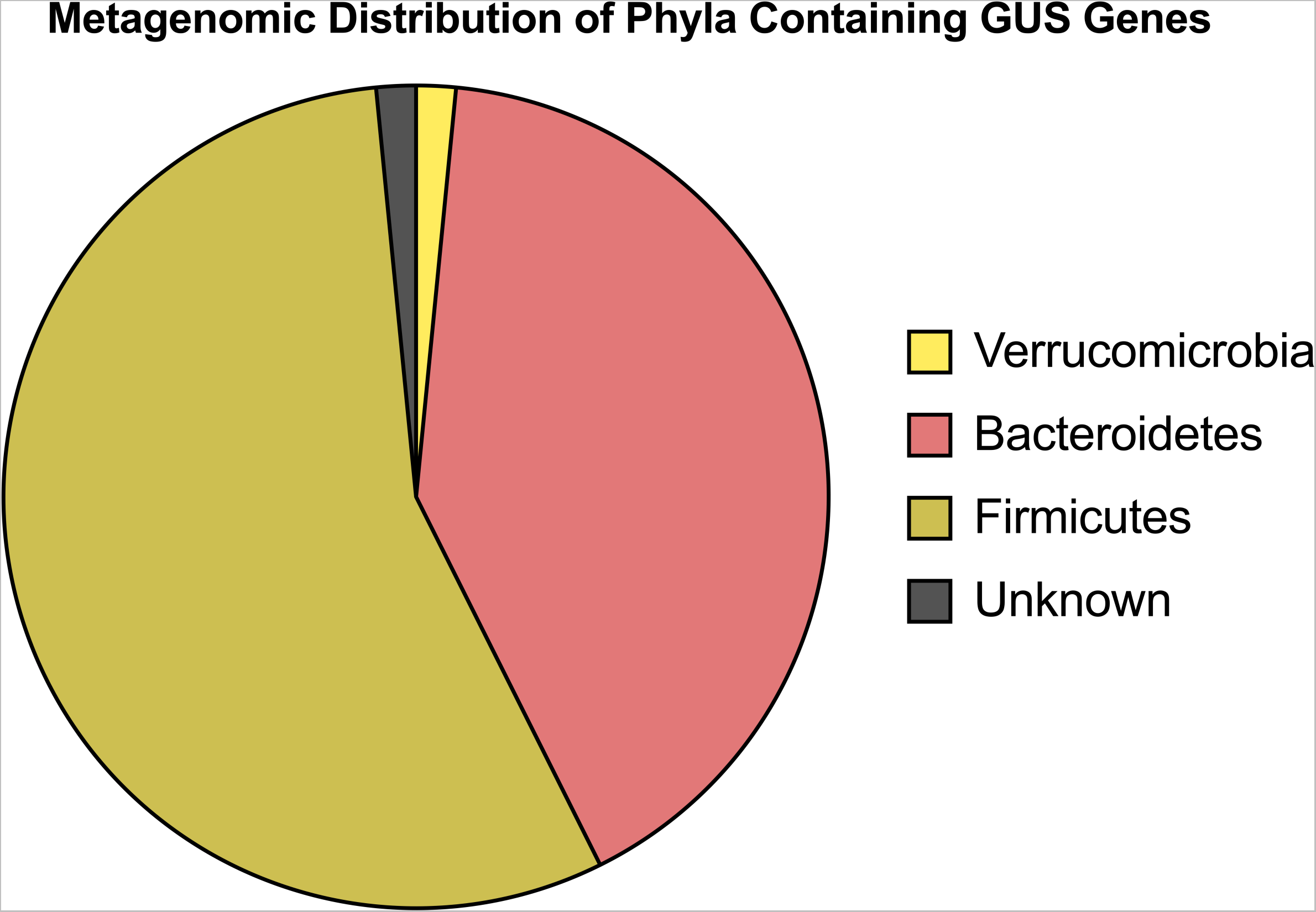
Taxonomic distribution of metagenome-derived GUS gene sequences by Phyla.

**Supplemental Figure 9.**
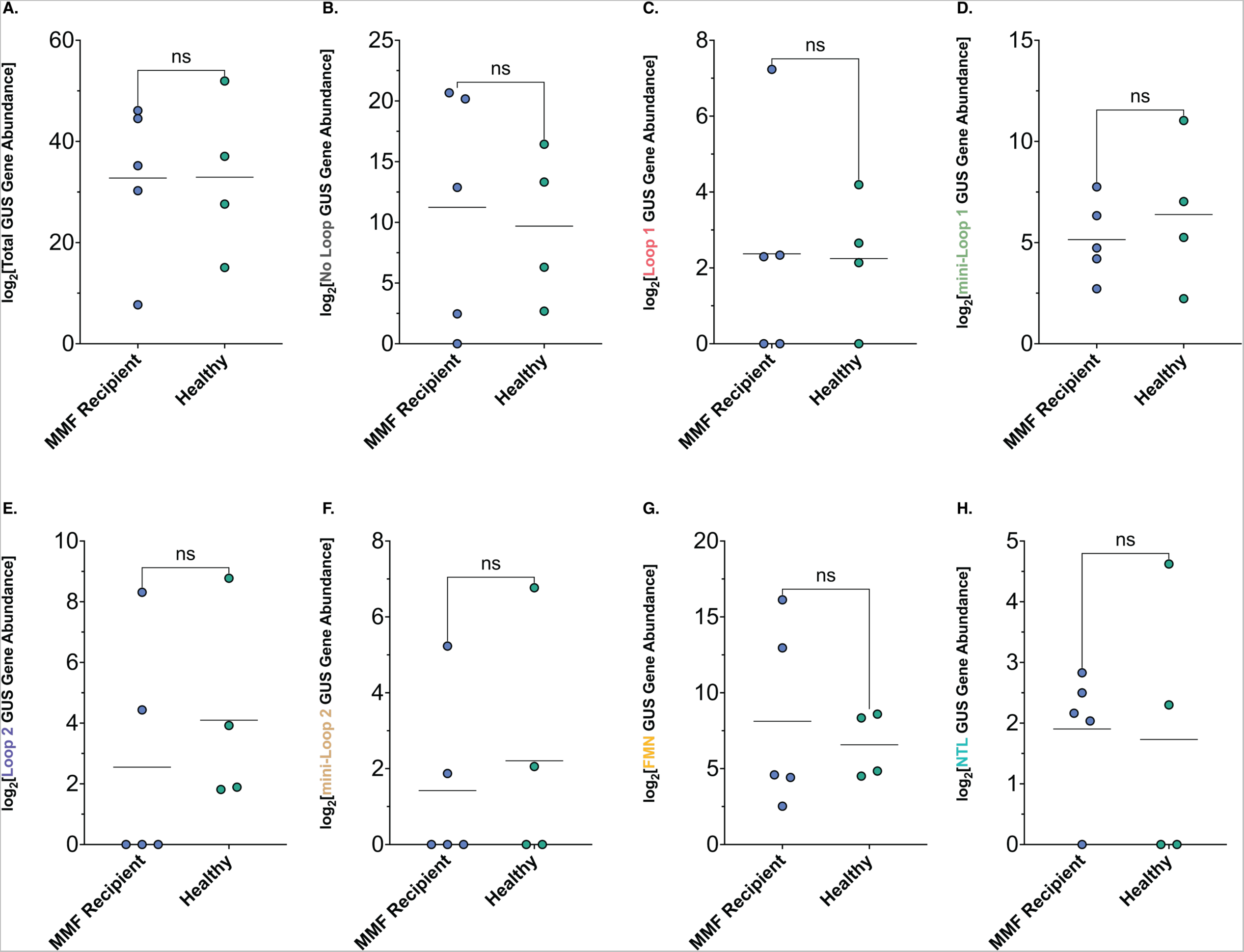
Comparison of normalized abundance of gene abundances for (A) Total GUS, (B) “No Loop” GUS, (C) “Loop 1” GUS, (D) “mini-Loop 1” GUS, (E) “Loop 2” GUS, (F) “mini-loop 2” GUS, (G) “FMN-binding” GUS, and (H) “N-Terminal Loop” GUS. Values were compared with Welch’s t-test; no significant correlations were present for any group.

**Supplemental Figure 10.**
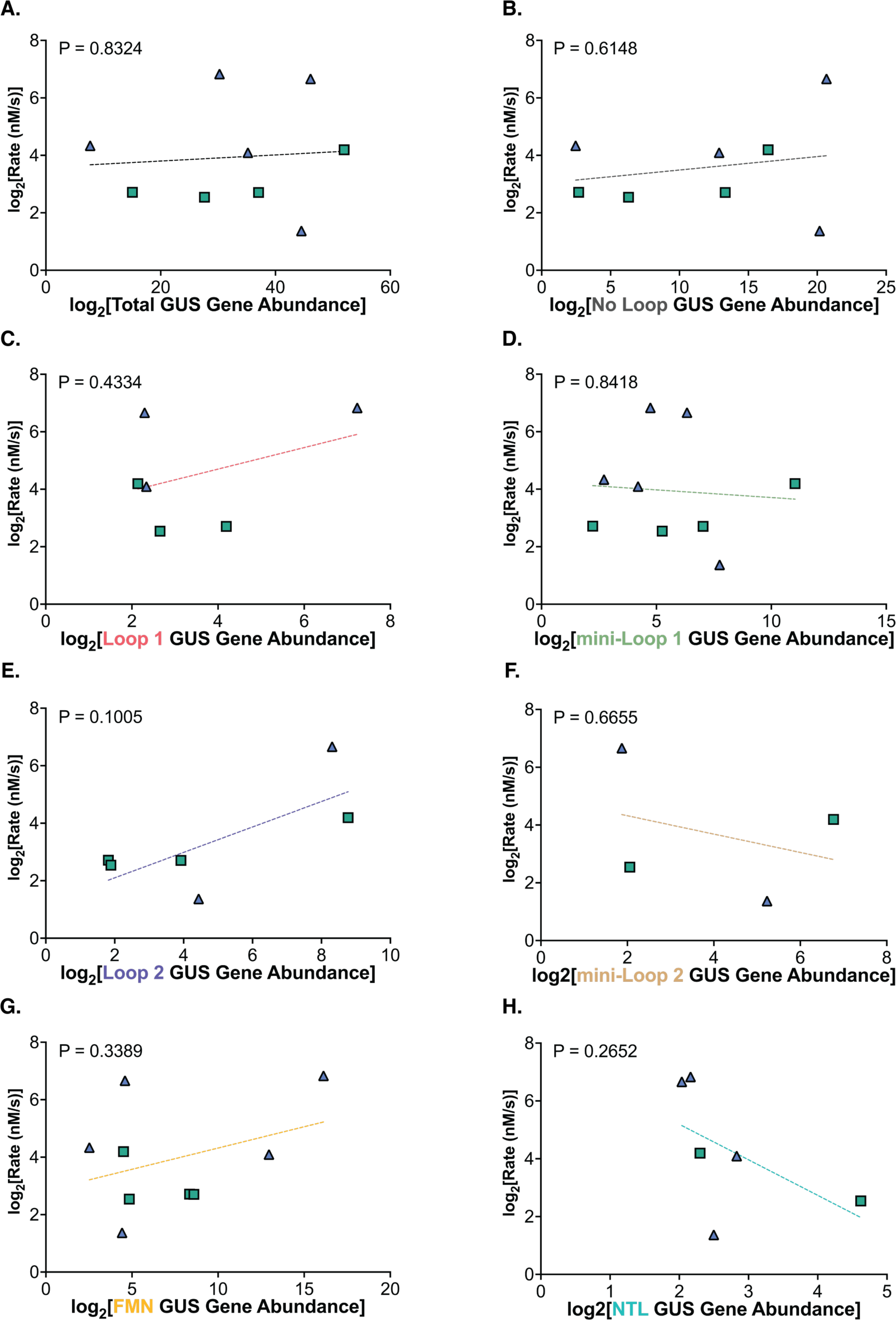
Correlation analysis between normalized rate of MPAG reactivation and normalized abundance of gene abundances for (A) Total GUS, (B) “No Loop” GUS, (C) “Loop 1” GUS, (D) “mini-Loop 1” GUS, (E) “Loop 2” GUS, (F) “mini-loop 2” GUS, (G) “FMN-binding” GUS, and (H) “N-Terminal Loop” GUS. P values reflect confidence in a slope that is significantly non-zero; no significant correlations were present for any group.

**Supplemental Figure 11.**
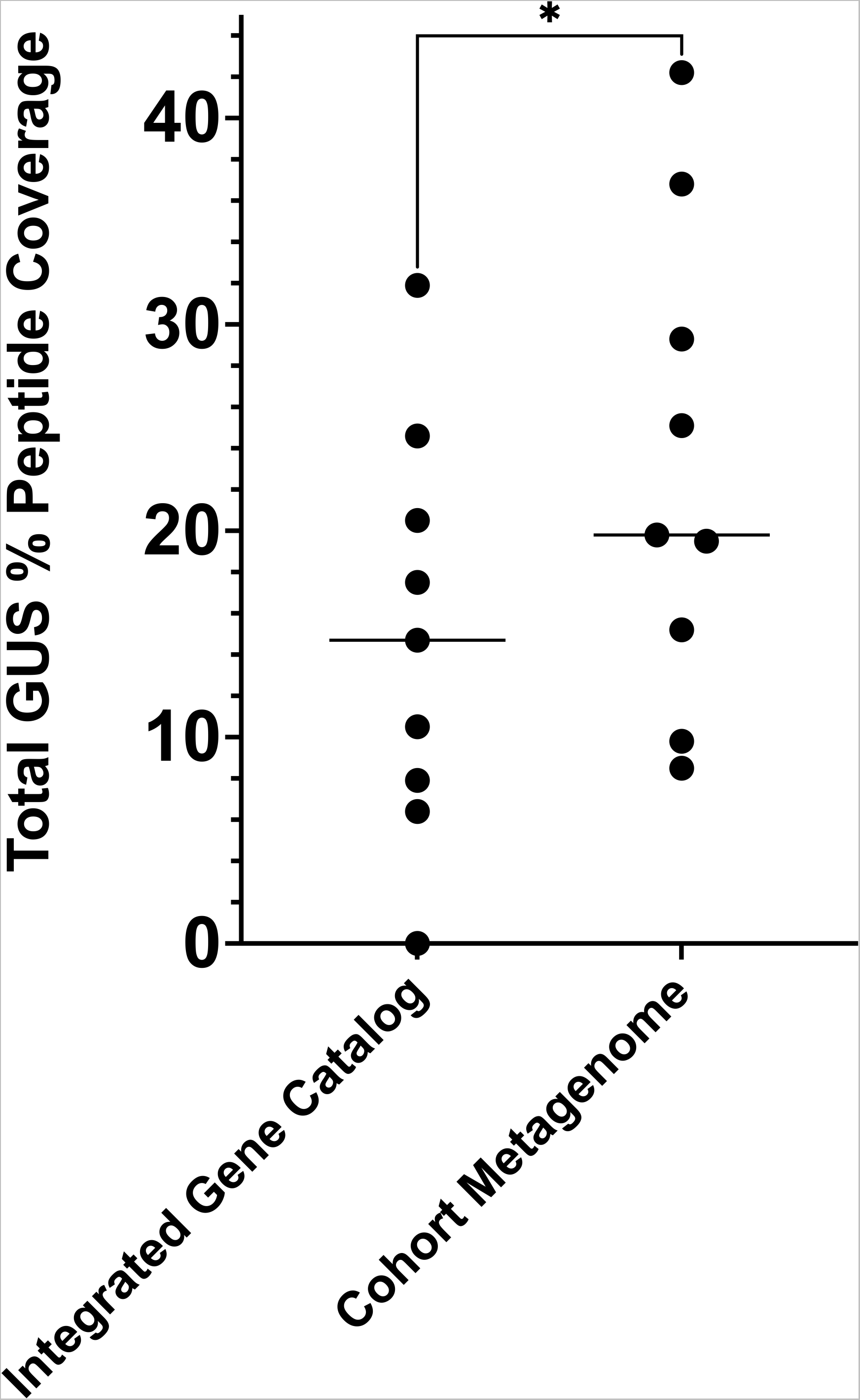
Total percent GUS peptide coverage when metaproteomics data are mapped to either the Integrated Gene Catalog or protein sequences derived from shotgun metagenomics performed specifically on the samples studied. P = 0.0312 when data is analyzed by a Wilcoxon two-tailed t-test.

**Supplemental Figure 12.**
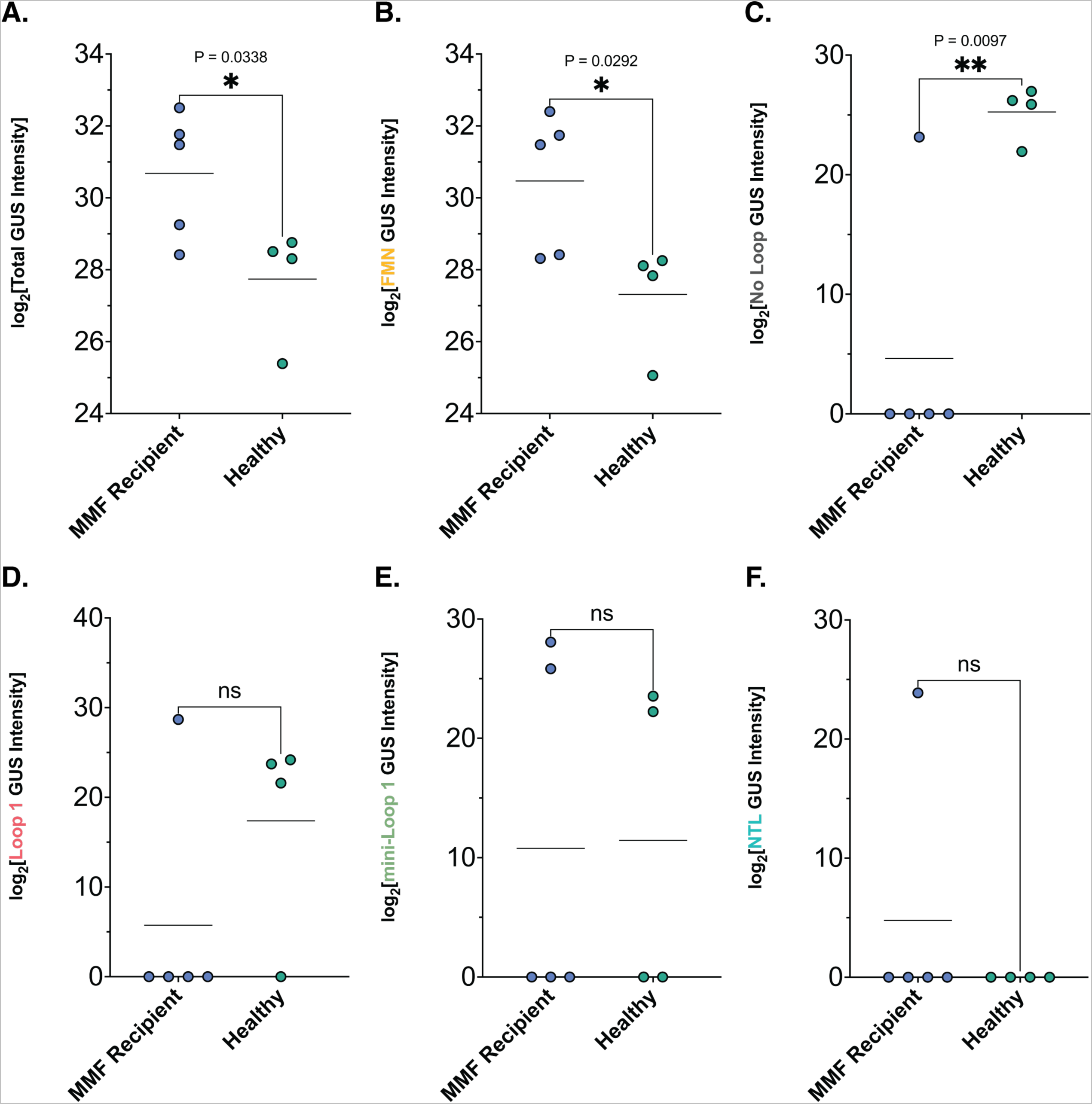
Comparison of normalized GUS abundance between treatment groups as determined by metaproteomics for (A) Total GUS, (B) “FMN-binding” GUS, (C) “No Loop” GUS, (D) “Loop 1” GUS, (E) “mini-Loop 1” GUS, and (F) “N-terminal Loop” GUS. Structural classes not shown here were not detected using metaproteomics. P values reflect comparison of normalized abundances using Welch’s t-test.

**Supplemental Figure 13.**
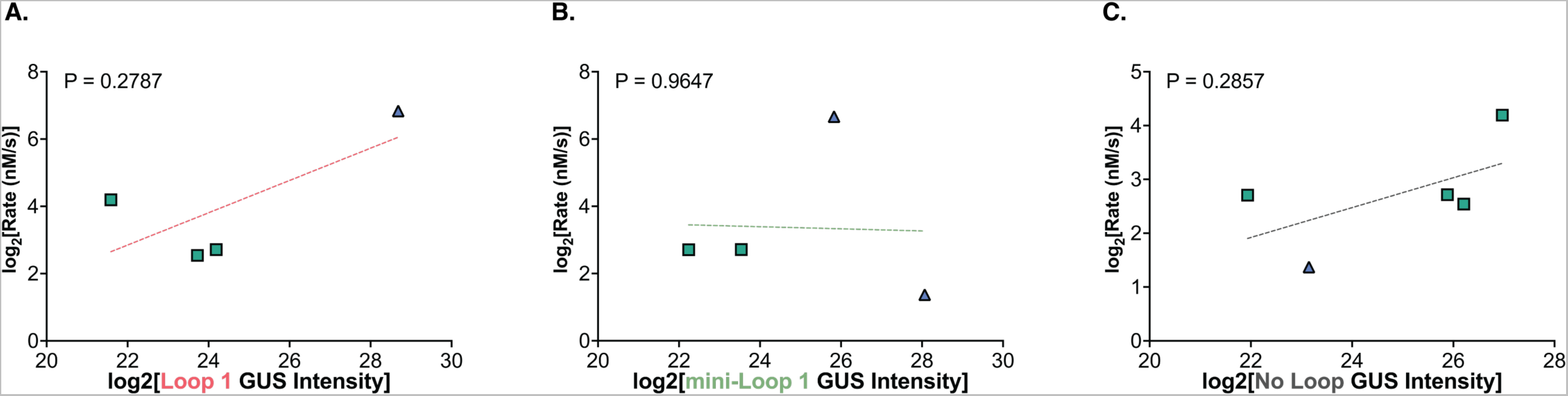
Correlation analysis between normalized rate of MPAG reactivation and normalized GUS Intensity for (A) “Loop 1” GUS, (B) “mini-Loop 1” GUS, and (C) “No Loop” GUS. Total GUS and FMN-binding GUS correlations are shown in Figure 5 D and E, respectively. Structural classes not shown here or in Figure 4 were not detected in more than 3 samples using metaproteomics. P values reflect confidence in a slope that is significantly non-zero; no significant correlations were present for any group shown in supplemental data.

**Supplemental Figure 14.**
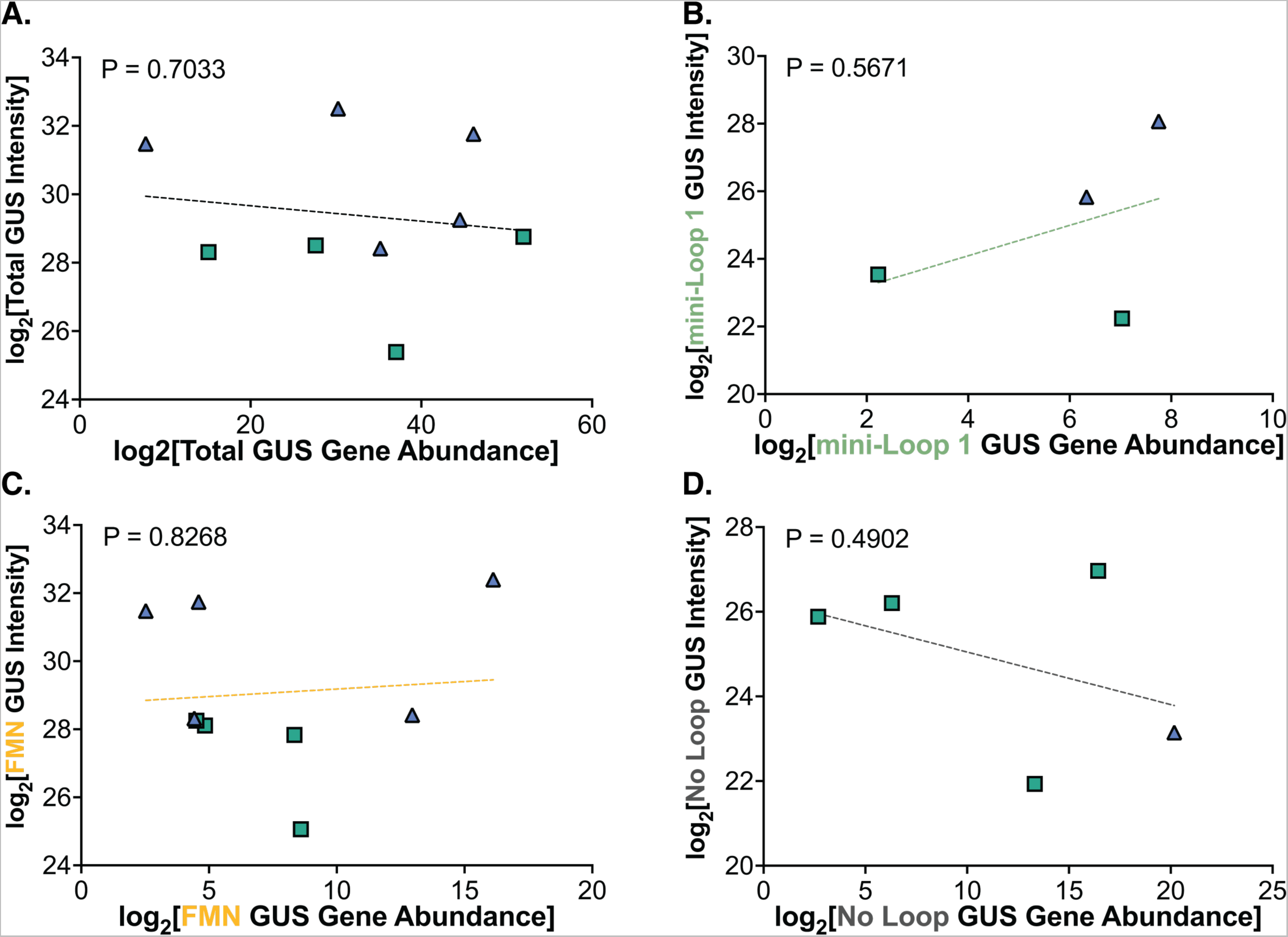
Correlation analysis between normalized (metagenomic) GUS gene abundance (x-axis) and normalized (metaproteomic) GUS Intensity for (**A**) Total GUS, (**B**) “mini-loop 1” GUS, (**C**) “FMN-binding” GUS, and (**D**) “No Loop” GUS. Structural classes not shown here were not detected in more than 3 samples between metagenomics and metaproteomics. P values reflect confidence in a slope that is significantly non-zero; no significant correlations were present for any group.

**Supplemental Table 1.** Cohort metadata.

**Supplemental Table 2.** Cohort metagenomics data.

**Supplementary Table 3.** Cohort metaproteomics data.

**Supplementary Table 4.** Statistics for regressions comparing relative percent abundance at every taxonomic level and rate of MPA reactivation for each donor. P value reflects confidence in a slope that is significantly non-zero.

## References

1. Park H, del Rosso J. 2011. The Emergence of Mycophenolate Mofetil in Dermatology. The Journal of Clinical and Aesthetic Dermatology. 4(1):18–27.

2. Allison AC, Eugui EM. 1993. The design and development of an immunosuppressive drug, mycophenolate mofetil. Springer Semin Immunopathology. 14:353–380.

3. Allison A. 2005. Mechanisms of action of mycophenolate mofetil. Lupus. 14:s2–s8. doi:10.1191/0961203305lu2109oa

4. Behrend M. 2001. Adverse Gastrointestinal Effects of Mycophenolate Mofetil. Drug Safety. 24(9):645–663.

5. Calmet FH, Yarur AJ, Pukazhendhi G, Ahmad J, Bhamidimarri KR. 2015. Endoscopic and histological features of mycophenolate mofetil colitis in patients after solid organ transplantation. Annals of Gastroenterology [Internet]. 28:366–373. www.annalsgastro.gr

6. Flannigan KL, Taylor MR, Pereira SK, Rodriguez-Arguello J, Moffat AW, Alston L, Wang X, Poon KK, Beck PL, Rioux KP, et al. 2018. An intact microbiota is required for the gastrointestinal toxicity of the immunosuppressant mycophenolate mofetil. Journal of Heart and Lung Transplantation. 37(9):1047–1059. doi:10.1016/j.healun.2018.05.002

7. Flannigan KL, Rajbar T, Moffat A, McKenzie LS, Dicke F, Rioux K, Workentine ML, Louie TJ, Hirota SA, Greenway SC. 2017. Changes in Composition of the Gut Bacterial Microbiome after Fecal Microbiota Transplantation for Recurrent Clostridium difficile Infection in a Pediatric Heart Transplant Patient. Frontiers in Cardiovascular Medicine. 4. doi:10.3389/fcvm.2017.00017

8. Taylor MR, Flannigan KL, Rahim H, Mohamud A, Lewis IA, Hirota SA, Greenway SC. 2019. Vancomycin Relieves Mycophenolate Mofetil-Induced Gastrointestinal Toxicity by Eliminating Gut Bacterial ß-Glucuronidase Activity. bioRxiv.:1–38. doi:org/10.1101/561274

9. Martin AM, Yabut JM, Choo JM, Page AJ, Sun EW, Jessup CF, Wesselingh SL, Khan WI, Rogers GB, Steinberg GR, Keating DJ. 2019. The gut microbiome regulates host glucose homeostasis via peripheral serotonin. Proc Natl Acad Sci U S A. 116(40):19802– 19804. doi:10.1073/pnas.1909311116

10. Koppel N, Rekdal VM, Balskus EP. 2017. Chemical transformation of xenobiotics by the human gut microbiota. Science (1979). 356(6344):1246–1257. doi:10.1126/science.aag2770

11. Chittim CL, Irwin SM, Balskus EP. 2018. Deciphering Human Gut Microbiota-Nutrient Interactions: A Role for Biochemistry. Biochemistry. 57(18):2567–2577. doi:10.1021/acs.biochem.7b01277

12. Redinbo MR. 2017. Microbial Molecules from the Multitudes within Us. Cell Metabolism [Internet]. 25(2):230–232. doi:10.1016/j.cmet.2017.01.013

13. Gilbert JA, Blaser MJ, Caporaso JG, Jansson JK, Lynch S V, Knight R. 2018. Current understanding of the human microbiome. Nat Med [Internet]. 24(4):392–400. doi:10.1038/nm.4517

14. Ervin SM, Redinbo MR. 2020. The Gut Microbiota Impact Cancer Etiology through “Phase IV Metabolism” of Xenobiotics and Endobiotics. Cancer Prev Res (Phila). 13(8):635–642. doi:10.1158/1940-6207.CAPR-20-0155

15. Lamba V, Sangkuhl K, Sanghavi K, Fish A, Altman RB, Klein TE. 2014. PharmGKB summary: mycophenolic acid pathway. Pharmacogenet Genomics. 24(1):73–79. doi:10.1097/FPC.0000000000000010

16. Nogueras F, Espinosa MD, Mansilla A, Torres JT, Cabrera MA, Martín-Vivaldi R. 2005. Mycophenolate mofetil-induced neutropenia in liver transplantation. In: Transplantation Proceedings. Vol. 37. [place unknown]; p. 1509–1511. doi:10.1016/j.transproceed.2005.02.038

17. Nguyen T, Park JY, Scudiere JR, Montgomery E. 2009. Mycophenolic Acid (Cellcept and Myofortic) Induced Injury of the Upper GI Tract [Internet]. [place unknown]. www.ajsp.com

18. Jardou M, Provost Q, Brossier C, Pinault É, Sauvage FL, Lawson R. 2021. Alteration of the gut microbiome in mycophenolate-induced enteropathy: impacts on the profile of short-chain fatty acids in a mouse model. BMC Pharmacology and Toxicology. 22(1). doi:10.1186/s40360-021-00536-4

19. Wallace BD, Wang H, Lane KT, Scott JE, Orans J, Koo JS, Venkatesh M, Jobin C, Yeh L, Mani S, Redinbo MR. 2010. Alleviating Cancer Drug Toxicity by Inhibiting a Bacterial Enzyme. Science (1979). 330:831–836. doi:10.1126/science.1191175

20. Ervin SM, Hanley RP, Lim L, Walton WG, Pearce KH, Bhatt AP, James LI, Redinbo MR. 2019. Targeting Regorafenib-Induced Toxicity through Inhibition of Gut Microbial β- Glucuronidases. ACS Chemical Biology. 14(12):2737–2744. doi:10.1021/acschembio.9b00663

21. Zhang J, Walker ME, Sanidad KZ, Zhang H, Liang Y, Zhao E, Chacon-Vargas K, Yeliseyev V, Parsonnet J, Haggerty TD, et al. 2022. Microbial enzymes induce colitis by reactivating triclosan in the mouse gastrointestinal tract. Nature Communications. 13(1). doi:10.1038/s41467-021-27762-y

22. Ervin SM, Li H, Lim L, Roberts LR, Liang X, Mani S, Redinbo MR. 2019. Gut microbial β-glucuronidases reactivate estrogens as components of the estrobolome that reactivate estrogens. Journal of Biological Chemistry. 294(49):18586–18599. doi:10.1074/jbc.RA119.010950

23. Jariwala PB, Pellock SJ, Cloer EW, Artola M, Simpson JB, Bhatt AP, Walton WG, Roberts LR, Davies GJ, Overkleeft HS, Redinbo MR. 2020. Discovering the Microbial Enzymes Driving Drug Toxicity with Activity-Based Protein Profiling. ACS Chemical Biology. 15(1):217–225. doi:10.1021/acschembio.9b00788

24. Pollet RM, D’Agostino EH, Walton WG, Xu Y, Little MS, Biernat KA, Pellock SJ, Patterson LM, Creekmore BC, Isenberg HN, et al. 2017. An Atlas of β-Glucuronidases in the Human Intestinal Microbiome. Structure [Internet]. 25(7):967–977.e5. doi:10.1016/j.str.2017.05.003

25. Pellock SJ, Walton WG, Biernat KA, Torres-rivera D, Creekmore BC, Xu Y, Liu J, Tripathy A, Stewart LJ, Redinbo MR. 2018. Three structurally and functionally distinct glucuronidases from the human gut microbe Bacteroides uniformis. JBC. 293:18559– 18573. doi:10.1074/jbc.RA118.005414

26. Pellock SJ, Walton WG, Ervin SM, Torres-Rivera D, Creekmore BC, Bergan G, Dunn ZD, Li B, Tripathy A, Redinbo MR. 2019. Discovery and Characterization of FMN-Binding β-Glucuronidases in the Human Gut Microbiome. Journal of Molecular Biology. 431(5):970–980. doi:10.1016/j.jmb.2019.01.013

27. Wu L, Jiang J, Jin Y, Kallemeijn WW, Kuo CL, Artola M, Dai W, van Elk C, van Eijk M, van der Marel GA, et al. 2017. Activity-based probes for functional interrogation of retaining β-glucuronidases. Nature Chemical Biology. 13(8):867–873. doi:10.1038/nchembio.2395

28. Li J, Jia H, Cai X, Zhong H, Feng Q, Sunagawa S, Arumugam M, Kultima JR, Prifti E, Nielsen T, et al. 2014. An integrated catalog of reference genes in the human gut microbiome. Nature Biotechnology. 32(8):834–841. doi:10.1038/nbt.2942

29. Jumper J, Evans R, Pritzel A, Green T, Figurnov M, Ronneberger O, Tunyasuvunakool K, Bates R, Žídek A, Potapenko A, et al. 2021. Highly accurate protein structure prediction with AlphaFold. Nature [Internet]. doi:10.1038/s41586-021-03819-2

30. Kaminski J, Gibson MK, Franzosa EA, Segata N, Dantas G, Huttenhower C. 2015. High-Specificity Targeted Functional Profiling in Microbial Communities with ShortBRED. PLoS Computational Biology. 11(12). doi:10.1371/journal.pcbi.1004557

31. Qian Y, Yang X, Xu S, Huang P, Li B, Du J, He Y, Su B, Xu LM, Wang L, et al. 2020. Gut metagenomics-derived genes as potential biomarkers of Parkinson’s disease. Brain. 143(8):2474–2489. doi:10.1093/brain/awaa201

32. Pacwa-Płociniczak M, Biniecka P, Bondarczuk K, Piotrowska-Seget Z. 2020. Metagenomic Functional Profiling Reveals Differences in Bacterial Composition and Function During Bioaugmentation of Aged Petroleum-Contaminated Soil. Frontiers in Microbiology. 11. doi:10.3389/fmicb.2020.02106

33. Nayfach S, Pollard KS. 2016. Toward Accurate and Quantitative Comparative Metagenomics. Cell. 166(5):1103–1116. doi:10.1016/j.cell.2016.08.007

34. Jariwala PB, Pellock SJ, Goldfarb D, Cloer EW, Artola M, Simpson JB, Bhatt AP, Walton WG, Roberts LR, Major B, et al. 2019. Discovering the Microbial Enzymes Driving Drug Toxicity with Activity-Based Protein Profiling. doi:10.1021/acschembio.9b00788

35. Robles-Vera I, de la Visitación N, Toral M, Sánchez M, Gómez-Guzmán M, Jiménez R, Romero M, Duarte J. 2021. Mycophenolate mediated remodeling of gut microbiota and improvement of gut-brain axis in spontaneously hypertensive rats. Biomedicine and Pharmacotherapy. 135. doi:10.1016/j.biopha.2020.111189

36. Yatsunenko T, Rey FE, Manary MJ, Trehan I, Dominguez-Bello MG, Contreras M, Magris M, Hidalgo G, Baldassano RN, Anokhin AP, et al. 2012. Human gut microbiome viewed across age and geography. Nature. 486(7402):222–227. doi:10.1038/nature11053

37. Pasolli E, Asnicar F, Manara S, Zolfo M, Karcher N, Armanini F, Beghini F, Manghi P, Tett A, Ghensi P, et al. 2019. Extensive Unexplored Human Microbiome Diversity Revealed by Over 150,000 Genomes from Metagenomes Spanning Age, Geography, and Lifestyle. Cell. 176(3):649–662.e20. doi:10.1016/j.cell.2019.01.001

38. Almeida A, Mitchell AL, Boland M, Forster SC, Gloor GB, Tarkowska A, Lawley TD, Finn RD. 2019. A new genomic blueprint of the human gut microbiota. Nature. 568(7753):499–504. doi:10.1038/s41586-019-0965-1

39. Falony G, Joossens M, Vieira-Silva S, Wang J, Darzi Y, Faust K, Kurilshikov A, Bonder MJ, Valles-Colomer M, Vandeputte D, et al. Population-level analysis of gut microbiome variation [Internet]. [place unknown]. https://www.science.org

40. Kultima JR, Coelho LP, Forslund K, Huerta-Cepas J, Li SS, Driessen M, Voigt AY, Zeller G, Sunagawa S, Bork P. 2016. MOCAT2: A metagenomic assembly, annotation and profiling framework. Bioinformatics. 32(16):2520–2523. doi:10.1093/bioinformatics/btw183

41. Beghini F, McIver LJ, Blanco-Míguez A, Dubois L, Asnicar F, Maharjan S, Mailyan A, Manghi P, Scholz M, Thomas AM, et al. 2021. Integrating taxonomic, functional, and strain-level profiling of diverse microbial communities with biobakery 3. Elife. 10. doi:10.7554/eLife.65088

42. Wickham H. 2016. ggplot2: Elegant Graphics for Data Analysis. [place unknown]: Springer-Verlag New York.

43. Bolyen E, Rideout JR, Dillon MR, et al. 2019. Reproducible, interactive, scalable and extensible microbiome data science using QIIME 2. Nature Biotechnology. 37(8):850– 852. doi:10.1038/s41587-019-0190-3

44. Sorgen A, Johnson J, Lambirth K, Clinton SM, Redmond M, Fodor A, Gibas C. 2021. Characterization of environmental and cultivable antibiotic-resistant microbial communities associated with wastewater treatment. Antibiotics. 10(4). doi:10.3390/antibiotics10040352

45. Langmead B, Salzberg SL. 2012. Fast gapped-read alignment with Bowtie 2. Nature Methods. 9(4):357–359. doi:10.1038/nmeth.1923

46. Hyatt D, Chen G-L, Locascio PF, Land ML, Larimer FW, Hauser LJ. 2010. Prodigal: prokaryotic gene recognition and translation initiation site identification [Internet]. [place unknown]. http://www.biomedcentral.com/1471-2105/11/119

47. Liao Y, Smyth GK, Shi W. 2014. FeatureCounts: An efficient general purpose program for assigning sequence reads to genomic features. Bioinformatics. 30(7):923–930. doi:10.1093/bioinformatics/btt656

48. Pereira MB, Wallroth M, Jonsson V, Kristiansson E. 2018. Comparison of normalization methods for the analysis of metagenomic gene abundance data. BMC Genomics. 19(1). doi:10.1186/s12864-018-4637-6

49. Camacho C, Coulouris G, Avagyan V, Ma N, Papadopoulos J, Bealer K, Madden TL. 2009. BLAST+: Architecture and applications. BMC Bioinformatics. 10. doi:10.1186/1471-2105-10-421

50. Li W, Godzik A. 2006. Cd-hit: A fast program for clustering and comparing large sets of protein or nucleotide sequences. Bioinformatics. 22(13):1658–1659. doi:10.1093/bioinformatics/btl158

51. Yu G, Smith DK, Zhu H, Guan Y, Lam TTY. 2017. ggtree: an r package for visualization and annotation of phylogenetic trees with their covariates and other associated data. Methods in Ecology and Evolution. 8(1):28–36. doi:10.1111/2041-210X.12628

52. Kanz C, Aldebert P, Althorpe N, Baker W, Baldwin A, Bates K, Browne P, van den Broek A, Castro M, Cochrane G, et al. 2005. The EMBL nucleotide sequence database. Nucleic Acids Research. 33(DATABASE ISS.). doi:10.1093/nar/gki098

53. Youn JY, Dunham WH, Hong SJ, Knight JDR, Bashkurov M, Chen GI, Bagci H, Rathod B, MacLeod G, Eng SWM, et al. 2018. High-Density Proximity Mapping Reveals the Subcellular Organization of mRNA-Associated Granules and Bodies. Molecular Cell. 69(3):517–532.e11. doi:10.1016/j.molcel.2017.12.020

54. Tyanova S, Temu T, Cox J. 2016. The MaxQuant computational platform for mass spectrometry-based shotgun proteomics. Nature Protocols. 11(12):2301–2319. doi:10.1038/nprot.2016.136

55. Bateman A, Martin MJ, Orchard S, Magrane M, Agivetova R, Ahmad S, Alpi E, Bowler-Barnett EH, Britto R, Bursteinas B, et al. 2021. UniProt: the universal protein knowledgebase in 2021. Nucleic Acids Research. 49(D1):D480–D489. doi:10.1093/nar/gkaa1100

56. Pellock SJ, Creekmore BC, Walton WG, Mehta N, Biernat KA, Cesmat AP, Ariyarathna Y, Dunn ZD, Li B, Jin J, et al. 2018. Gut Microbial β-Glucuronidase Inhibition via Catalytic Cycle Interception. ACS Central Science. 4(7):868–879. doi:10.1021/acscentsci.8b00239

57. Mailman MD, Feolo M, Jin Y, Kimura M, Tryka K, Bagoutdinov R, Hao L, Kiang A, Paschall J, Phan L, et al. The NCBI dbGaP database of genotypes and phenotypes [Internet]. [place unknown]. www.ncbi.nlm.nih.gov/projects/gap/pdf/dbgap_2b_security_procedures.pdf.

58. Chen IMA, Chu K, Palaniappan K, Pillay M, Ratner A, Huang J, Huntemann M, Varghese N, White JR, Seshadri R, et al. 2019. IMG/M v.5.0: An integrated data management and comparative analysis system for microbial genomes and microbiomes. Nucleic Acids Research. 47(D1):D666–D677. doi:10.1093/nar/gky901

59. Vizcaíno JA, Csordas A, Del-Toro N, Dianes JA, Griss J, Lavidas I, Mayer G, Perez-Riverol Y, Reisinger F, Ternent T, et al. 2016. 2016 update of the PRIDE database and its related tools. Nucleic Acids Research. 44(D1):D447–D456. doi:10.1093/nar/gkv1145

